# A multi-component, community-based strategy to facilitate COVID-19 vaccine uptake among Latinx populations: from theory to practice

**DOI:** 10.1101/2021.06.07.21258230

**Authors:** Carina Marquez, Andrew D. Kerkhoff, Jamie Naso, Maria G. Contreras, Edgar Castellanos, Susana Rojas, James Peng, Luis Rubio, Diane Jones, Jon Jacobo, Susy Rojas, Rafael Gonzalez, Jonathan D. Fuchs, Douglas Black, Salustiano Ribeiro, Jen Nossokoff, Valerie Tulier-Laiwa, Jacqueline Martinez, Gabriel Chamie, Genay Pilarowski, Joseph DeRisi, Maya Petersen, Diane V. Havlir

## Abstract

**Background:** COVID-19 vaccine coverage in the Latinx community depends on delivery systems that overcome barriers such as institutional distrust, misinformation, and access to care. We hypothesized that a community-centered vaccination strategy that included mobilization, vaccination, and “activation” components could successfully reach an underserved Latinx population, utilizing its social networks to boost vaccination coverage.

**Methods and Findings:** Our community-academic-public health partnership, “Unidos en Salud,” utilized a theory-informed approach to design our “Motivate, Vaccinate, and Activate” COVID-19 vaccination strategy. Our strategy’s design was guided by the PRECEDE Model and sought to address and overcome predisposing, enabling, and reinforcing barriers to COVID-19 vaccination faced by Latinx individuals in San Francisco. We evaluated our prototype outdoor, “neighborhood” vaccination program located in a central commercial and transport hub in the Mission District in San Francisco, using the Reach, Effectiveness, Adoption, Implementation and Maintenance (RE-AIM) framework during a 16-week period from February 1, 2021 to May 19, 2021. Programmatic data, city-wide COVID-19 surveillance data, and a survey conducted between May 2, 2021 and May 19, 2021 among 997 vaccinated clients ≥16 years old were used in the evaluation.

There were 20,792 COVID-19 vaccinations administered at the neighborhood site during the 16-week evaluation period. Vaccine recipients had a median age of 43 (IQR 32-56) years, 53.9% were male and 70.5% were Latinx, 14.1% white, 7.7% Asian, 2.4% Black, and 5.3% other. Latinx vaccinated clients were substantially more likely than non-Latinx clients to have an annual household income of less than $50,000 a year (76.1% vs. 33.5%), be a first-generation immigrant (60.2% vs. 30.1%), not have health insurance (47.3% vs. 16.0%), and not have access to primary care provider (62.4% vs. 36.2%). The most frequently reported reasons for choosing vaccination at the site were its neighborhood location (28.6%), easy and convenient scheduling (26.9%) and recommendation by someone they trusted (18.1%); approximately 99% reported having an overall positive experience, regardless of ethnicity. Notably, 58.3% of clients reported that they were able to get vaccinated earlier because of the neighborhood vaccination site, 98.4% of clients completed both vaccine doses, and 90.7% said that they were more likely to recommend COVID-19 vaccination to family and friends after their experience; these findings did not substantially differ according to ethnicity. There were 40.3% of vaccinated clients who said they still knew at least one unvaccinated person (64.6% knew ≥3). Among clients who received both vaccine doses (n=729), 91.0% said that after their vaccination experience, they had personally reached out to at least one unvaccinated person they knew (61.6% reached out to ≥3) to recommend getting vaccinated; 83.0% of clients reported that one or more friends, and/or family members got vaccinated as a result of their outreach, including 18.9% who reported 6 or more persons got vaccinated as a result of their influence.

**Conclusions:** A multi-component, “Motivate, Vaccinate, and Activate” community-based strategy addressing barriers to COVID-19 vaccination for the Latinx population reached the intended population, and vaccinated individuals served as ambassadors to recruit other friends and family members to get vaccinated.

## Introduction

COVID-19 has disproportionately affected underserved communities of color, including Latinx in the United States, further amplifying long-standing health disparities [1–4]. The highly safe and effective COVID-19 vaccines are the most critical tool in our public health strategy to overcome the COVID-19 pandemic. The success of our public health vaccination strategy relies on our ability to rapidly reach the population at highest risks of COVID-19, including communities of color which have been subject to decades of inequities and often have the least access to health care, including vaccination [5, 6].

There is a growing understanding of the barriers to vaccination communities of color face in the United States. In California where Latinx persons account for approximately 40% of the total population and 63% of COVID-19 cases to date, they have only received 27% of all COVID-19 vaccinations administered statewide [7, 8]. Some of the barriers to COVID-19 vaccine uptake include a lack of trust in health systems stemming from historical experience, structural racism, inaccurate and insufficient information, and structural barriers to vaccine access [2,9–11]. There are few formal evaluations of vaccination strategies that can overcome these barriers, and none for COVID-19 vaccines.

We hypothesized that a community-centered, culturally-tailored, theory-informed vaccination strategy that included mobilization, vaccination and “activation” components could successfully reach an underserved Latinx population, utilizing its social networks to boost vaccination coverage. We developed this multifaceted approach via a community-academic-public health partnership founded in April, 2020 to increase SARS-COV-2 testing and public health surveillance in the Mission neighborhood in San Francisco. In this paper we describe the “Motivate, Vaccinate, and Activate” vaccination program and evaluate the program according to the Reach, Effectiveness, Adoption, Implementation and Maintenance (RE-AIM) framework [12] during a 16-week period from February 1 and May 19^th^ 2021.

## Methods

### Setting

The Unidos en Salud (UeS) neighborhood vaccination program was implemented in the Mission District, home to a large Latinx and immigrant community in San Francisco, California [13]. The Mission Neighborhood comprises a large majority of the 94110 zip code, which has an estimated population of 72,380 persons (62,452 ≥16 years old) of whom 33.4% identify as Latinx, 43.8% white, 14.7% Asian and 3.3% Black [14]. The neighborhood is economically heterogenous; the median household income is $134,592 per year, yet 22.6% of households have a combined income less than $50,000. The Mission District is an important cultural and commercial hub for Latinx people living throughout the many neighborhoods of San Francisco’s Southeast sector, which have consistently had the highest rates of COVID-19 throughout the pandemic (**Supplementary Figure 1**), concentrated among low-income, front line workers, unable to work from home [1, 15]. UeS has offered free walk-up, COVID-19 testing in the Mission District since April, 2020.

### Ethics

The study was conducted under a public health surveillance program reviewed by UCSF Committee on Human Research. Survey participants provided consent in their preferred language prior to survey initiation.

### Community-academic-public health partnership model

Unidos en Salud (“United in Health”, UeS) is a community-academic-public health partnership founded in April 2020 to respond to and support the Latinx community in San Francisco during the COVID-19 pandemic. The partnership provides ongoing SARS-CoV-2 testing and surveillance [1,15–18]. The partnership includes the San Francisco Latino Task Force on COVID-19 (LTF), the University of California, San Francisco (UCSF), the University of California, Berkeley, the Chan Zuckerberg Biohub, locally owned Bay Area Phlebotomy and Laboratory Services (BayPLS), Primary.Health, and the San Francisco Department of Public Health (SFDPH). The LTF is a group made up of members and leaders from more than three dozen Latinx, community-based organizations, many of which are long-standing, that was forged during the COVID-19 pandemic [19]. Primary.Health informatics was founded in 2020 to meet community based COVID-19 testing efforts and provides cloud-based support for COVID-19 testing and vaccination registration and data metric tracking. BayPLS has bilingual staff that have provided community testing and vaccination services in the San Francisco Bay Area since the beginning of the COVID-19 pandemic. UeS operates via joint decision making by leaders from each of the partners and academic institution faculty that occur at weekly meetings. Funding for the vaccination program was provided by a combination of the SFDPH, UCSF, private donors and the Chan-Zuckerberg Initiative.

### Overview and design of a strategy to reach and increase COVID-19 vaccination among Latinx individuals

For our vaccine strategy prototype, we utilized a theory-informed approach to design a multicomponent, implementation strategy that addressed barriers to COVID-19 vaccination faced by Latinx and other community members (**Table 1**). We specifically sought to reach those community members for whom the City’s high volume vaccination sites posed barriers such as a lack of transportation and “institutional mistrust.” We built upon lessons learned providing community-based COVID-19 services to socioeconomically vulnerable individuals [13–15], including providing free walk-up SARS-CoV-2 testing, with support for persons testing positive, to over 30,000 persons since April 2020. Formative findings from this work were used to design our “Motivate, Vaccinate, and Activate” strategy (**Figure 1)**. The “Motivate, Vaccinate, and Activate” strategy’s design was guided by the PRECEDE Model [20]and therefore sought to address and overcome predisposing, enabling and reinforcing barriers to COVID-19 vaccination faced by Latinx and other low-income community members in San Francisco (**Table 1).** The “Motivate, Vaccinate, and Activate” strategy represents a culturally-tailored, multicomponent implementation strategy to optimize reach and uptake of COVID-19 vaccination among Latinx individuals in San Francisco. Further details for each strategy component are provided below and are summarized in **Table 1**.

**Figure 1.**
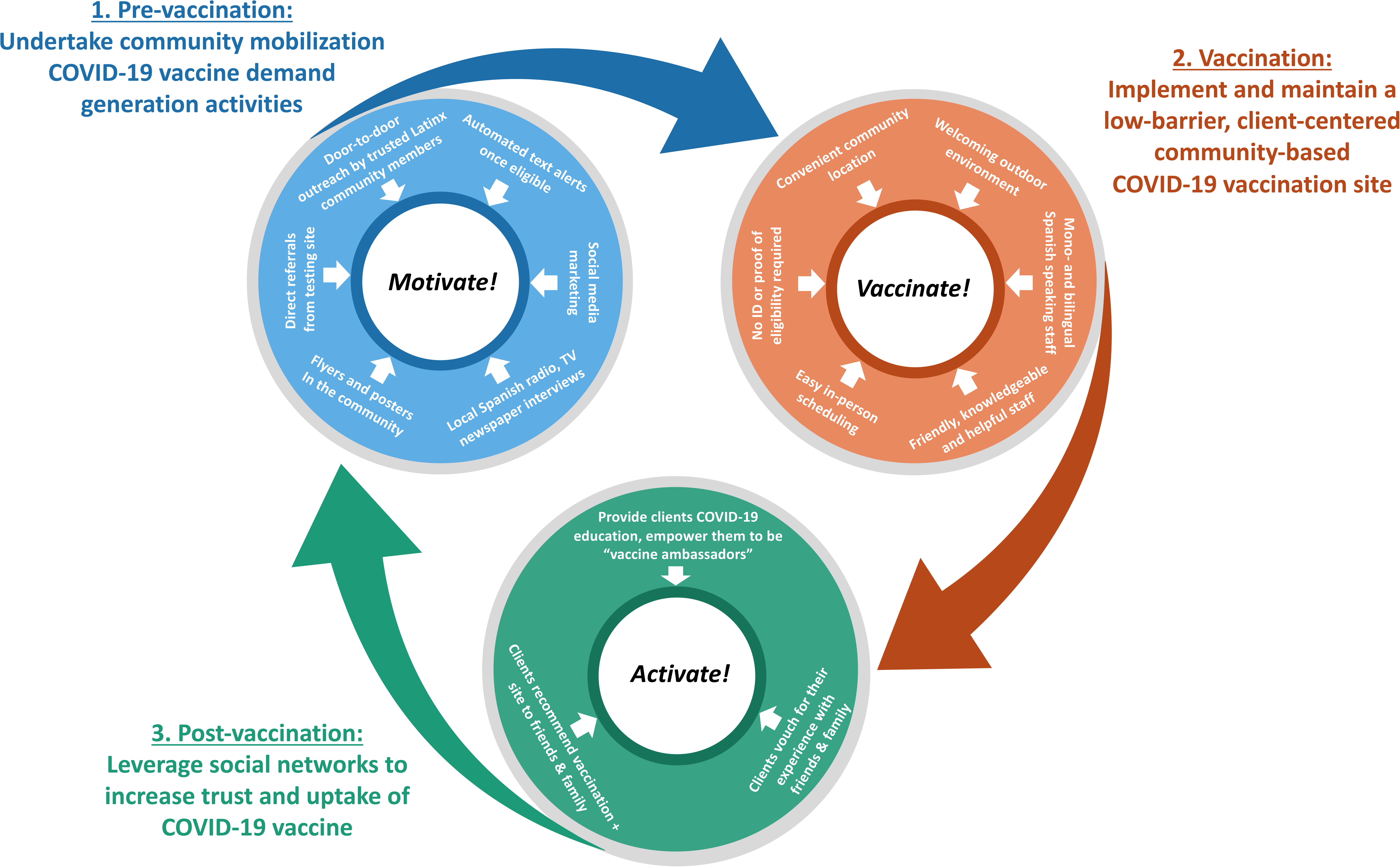
Overview of the “Motivate, Vaccinate and Activate” Strategy to increase vaccine uptake among Latinx community members living in San Francisco.

**Table 1.** Description of the “Motivate, Vaccinate, and Activate” Strategy to address predisposing, enabling and reinforcing factors to COVID-19 vaccination among low-income Latinx individuals.

| Influencing factors | Barriers to COVID-19 vaccination | Strategy component | Description of intervention activities that directly address barriers |
| --- | --- | --- | --- |
| **Predisposing**<br>(Knowledge, attitudes, beliefs, skills, values, self-efficacy) | Questions and concerns about vaccine safety and efficacy:<br>Vaccine efficacy and impact on transmission.<br>Short- and long-term side-effects and safety<br>Speed of development and approval process.<br>Trust:<br>Concerns about impact on public charge and immigration status.<br>Mistrust of healthcare systems or government.<br>Knowledge and awareness of eligibility requirements:<br>Unsure when eligible and whether proof of eligibility needed.<br>Questions about whether eligibility applied to people with comorbid conditions.<br>Questions about whether people with history of COVID-19 were eligible.<br>Unsure where to get the COVID-19 vaccine.<br>Availability of language concordant messaging and registration. | **Motivate**<br>(Community mobilization and demand generation activities) | Trusted and known bilingual or monolingual Spanish-speaking Latinx community members provided direct outreach, including going door-to-door to businesses and senior living facilities, giving interviews on Spanish language radio shows, and hosting vaccine townhalls to raise awareness about vaccine eligibility, provide vaccine-related education and raise awareness about the UeS vaccination site.<br>Community health workers provided face-to-face outreach and vaccine education hand-outs to people attending UeS neighborhood COVID-19 testing site starting one-month before the vaccination site opened.<br>Community-based organizations able to directly schedule eligible persons that they service (e.g., able to gather and submit list of older clients who wanted to receive a vaccine).<br>Sunday ‘Cafecitos’ across from vaccination site where community members could walk up and ask questions to Latinx physicians while enjoying pan dulce and coffee.<br>Text messages and site information sent to all clients who had ever been tested for COVID-19 at UeS community site as soon as they became eligible for COVID-19 vaccination.<br>Dissemination of information about benefits of vaccination, UeS vaccination site and eligibility via posters in the community, social media, community websites, local Spanish language radio stations and newspapers. These were updated throughout the implementation period as eligibility criteria changed.<br>Direct, vaccination referral from UeS COVID-19 testing site. |
|  |  | **Vaccinate**<br>(Community-based, low-barrier, client-centered, vaccination site) | Vaccination site co-developed and run in partnership between trusted community-led organization (LTF) and UCSF which has deep roots in the community, working hand-in-hand to provide COVID-19 services since April 2020.<br>All site staff provided COVID-19 vaccine-related education and regular refresher trainings to be able to answer any community member/client questions or address any concerns.<br>Site staff largely mono- or bilingual Spanish speaking and all trained and refreshed on good customer service principles. |
|  |  | **Activate**<br>(Leverage social networks to increase vaccine uptake) | Peer vaccine ambassadors speak with unvaccinated friends and family to provide COVID-19 vaccine, including its benefits, share their positive experiences at the neighborhood site, share information about how/where to register for vaccination at the neighborhood site, and recommend that they get vaccinated. |
| **Enabling**<br>(Availability and accessibility of resources, policies, laws) | No mobile phone/computer access to schedule vaccination appointments online.<br>Difficult to navigate online vaccination appointments.<br>Requirement to prove residency or vaccine eligibility for scheduling.<br>Requirement to prove eligibility.<br>No personal transportation.<br>Direct + indirect costs of time required to get scheduled and vaccinated (financial insecurity).<br>Lack of language/culturally-concordant messaging, registration and vaccination services.<br>No health insurance or not linked to primary healthcare services. | **Vaccinate**<br>(Community-based, low-barrier, client-centered, vaccination site) | Vaccination site is community-based and conveniently located near a busy transport hub – “in the neighborhood.”<br>Outdoor, welcoming environment with all site staff well-trained and refreshed on good customer service principles, emphasizing kindness and helpfulness.<br>Culturally-tailored site with bicultural, bilingual staff, many of whom are local community members.<br>Low-barrier scheduling features included:<br>On-site registration 7 days a week<br>Self-attestation of meeting eligibility requirements, no requirement to show an ID to prove residency, or to prove vaccine eligibility or health insurance status.<br>Sunday appointments offered.<br>Offering walk-up appointments over time.<br>Close collaboration with SFDPH to ensure consistent vaccine supply and avoid appointment cancellation. |
| **Reinforcing**<br>(Social support, influence of peers, family members, and general community attitudes) | Lack of friends, family members, and coworkers, who are vaccinated and can serve as credible source for vaccine-related knowledge and experiences.<br>Lack of friends and family members loved ones/peers encouraging vaccination through empathy and support. | **Activate**<br>(Leverage social networks to increase vaccine uptake) | Community health team members approach clients while waiting for 15-30 minutes <u>after</u> their vaccination.<br>Clients are counselled about side effects a person may expect.<br>Clients are then provided additional education about COVID-19 vaccines and any vaccine-related questions are answered.<br>Clients are then encouraged to reach out to unvaccinated family members and peers to provide COVID-19-related vaccine education, information about UeS vaccination site and personally recommend that they get vaccinated. |

### “Motivate, Vaccinate, and Activate” strategy components

#### Community mobilization and demand generation activities (“Motivate”)

We used several methods to inform community members about COVID-19 vaccination and its benefits, raise awareness about COVID-19 vaccine eligibility and the UeS neighborhood vaccination site, and ultimately increase motivation and demand for COVID-19 vaccination (**Table 1)**. Responses to a survey on vaccine attitudes and preferences from community members seeking COVID-19 testing at our site in January 2021 were used to inform training of our community workers and informational materials on vaccination [9]. Vaccine efficacy, short- and long-term side effects of vaccines, and conspiracy theories on motivation behind vaccine development were some of the key topics of concern that were directly addressed through educational and outreach efforts

UeS community workers performed direct community outreach in the Mission District, via door-to-door household canvassing with flyers and by speaking to business owners in the commercial corridor of the Mission. We also emphasized to elder care facilities the opportunity and rationale to vaccinate high-risk adults. The LTF reached out to their multiple CBO’s and network of community organization to push out invitations to priority groups such as community health workers as they became eligible; this guaranteed that we were reaching our key populations who didn’t necessarily have access to the mass media advertising of vaccination appointments. Additionally, automated text messages (and reminder texts) were sent to 26,206 unique phone numbers of community members who had previously been tested for COVID-19 at a UeS site as soon as they became eligible for vaccination and invited them to get vaccinated at the UeS neighborhood site. Furthermore, flyers and posters were posted throughout the community (**Figure 2a**), and UeS members undertook Spanish radio, newspaper and television interviews to feature the UeS neighborhood vaccination site. Community leaders vaccinated at the site posted photos on social media (Facebook, TikTok), encouraging others to get vaccinated. Additionally, to engage community members who trust physicians but do not have access to one, we hosted Sunday ‘cafecitos’ directly across from the vaccination site, where community members could walk up and ask Latinx physicians questions while enjoying free pan dulce and coffee.

**Figure 2.**
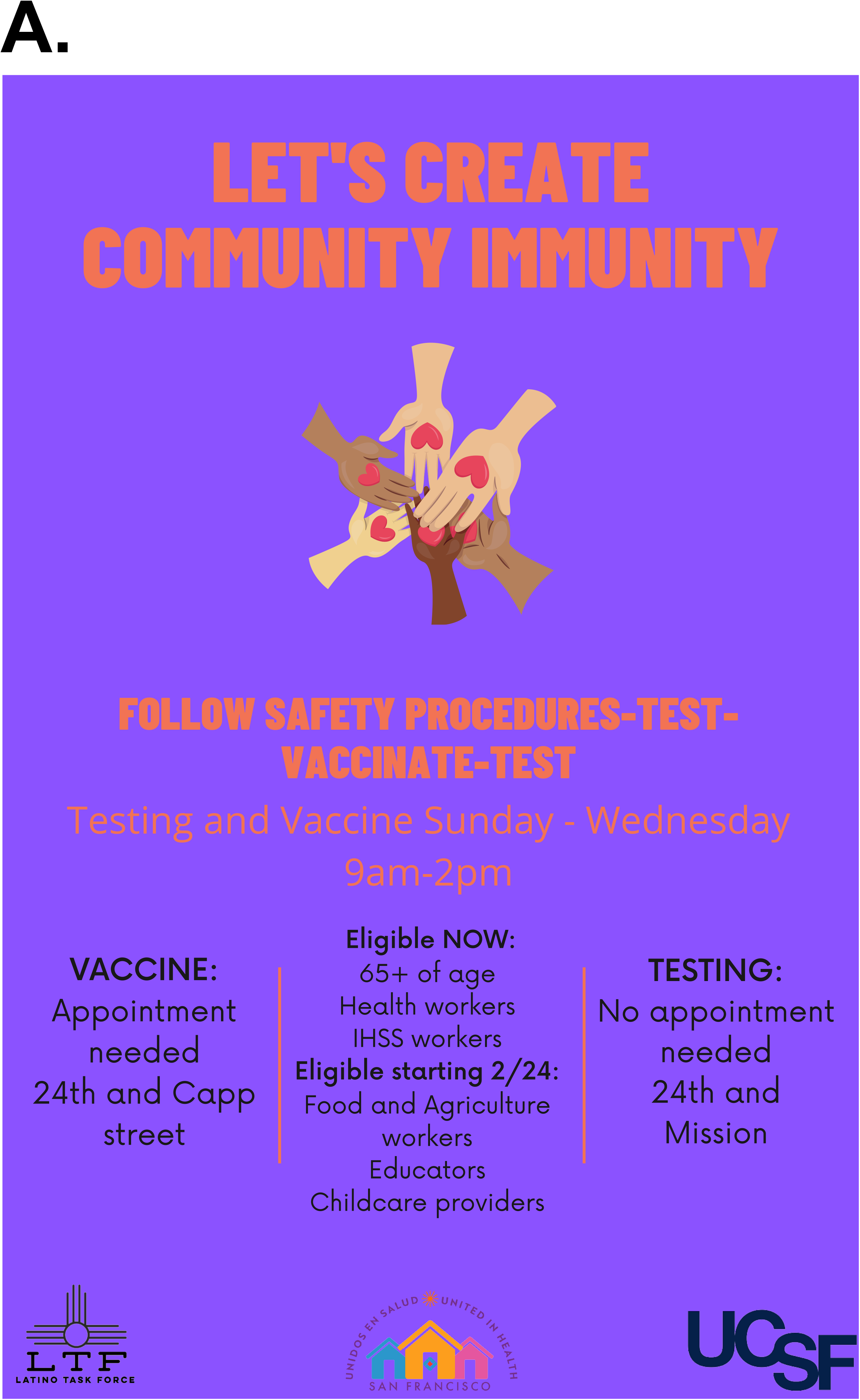
Photographs taken during the implementation period of the “Motivate, Vaccinate and Activate”. Panel A shows an example of flyers promoting COVID-19 vaccination at the Unidos en Salud neighborhood site, posted in English and Spanish. Panel B shows the registration area of the Unidos en Salud low-barrier vaccination site. Panel C shows the Unidos en Salud COVID-19 testing site directly across from the vaccination site. Community members could make vaccination appointments and were also encouraged to drop-in for same-day vaccination. Community members could ask site staff questions about the COVID-19 vaccine and were provided handouts about the COVID-19 vaccine. Panel D shows the waiting area of the Unidos en Salud low-barrier vaccination site where clients were provided education on possible COVID-19 side effects and were also ‘activated’ to the community health team be become vaccine ambassadors to reach out to their unvaccinated family members, friends and co-workers.

#### A client-centered neighborhood vaccination site (“Vaccinate”)

##### Neighborhood vaccination site characteristics

The UeS neighborhood vaccination site was the first of 8 community-led sites established in partnership with the SFDPH to increase equitable distribution of vaccines to neighborhoods disproportionately affected by COVID-19. The site was located outdoors and was located in a small parking lot (approximately 8,800 square feet), behind a McDonald’s restaurant at 24^th^ and Capp Street (**Figure 2b)**. The vaccination site was across the street from the free, walk-up UeS COVID-19 testing site, located at a busy public plaza and transport hub – the intersection of both above ground bus/streetcar system (MUNI) and underground subway system (BART) (**Figure 2c**). This location was intentionally chosen in order to enhance visibility and promote walk-up scheduling as people exited public transit and walked through the neighborhood. The site was open 4 days a week (Sunday through Wednesday) between the hours of 9am and 4pm. It was designed to be open in order to improve flow, and had several semi-permanent tents corresponding to different aspects of the vaccination process (i.e., check-in, pre-vaccination waiting area, vaccination area, and post-vaccination waiting area) (**Figure 2d**); the tents also provided privacy from the busy surrounding area as well as protection from the elements. The site played music in order to attract persons passing by and to create a welcoming and positive environment for those attending the site. Large, colorful signs in both Spanish and English were hung around and near the site to generate further awareness and encourage community members to register to get vaccinated.

The vaccine site officially opened on February 1, 2021 and remains operational. It provided clients either the Pfizer or Moderna COVID-19 mRNA two-dose vaccine, depending on availability. In order to minimize inconvenience for busy, socioeconomically vulnerable community members, the vaccine strategy prioritized smooth logistics and avoiding any need to reschedule appointments. To this end, while following dosage segment regulations we worked closely with the SFPDH and California health officials to mitigate any disruptions in vaccine availability. We further limited vaccination appointments to reduce the likelihood of vaccine stock-out and to facilitate site logistics. As we developed clinic operating protocols and improved efficiency, we were able to raise the appointment cap to 500 per day.

##### Vaccination site personnel and customer service principles (client-centeredness)

The UeS neighborhood vaccination site was predominantly staffed by trained members of the local community who were bicultural and bilingual or monolingual Spanish speakers. Vaccinations were provided by bicultural and bilingual Spanish-speaking BayPLS and UeS staff, many whom had worked for prior UeS mass community-based COVID-19 testing events. The number of staff on site changed throughout the implementation period based on demand and ranged between 25 and 30 and peaked at 40 during mid-April 2021 when the general population became eligible for the COVID-19 vaccine in California. Staffing at the site consisted of 6 persons registering clients for vaccination appointments, 3 persons greeting and checking-in clients, 12 people preparing and administering vaccines, 2 people assisting clients with translation and navigation, 2 people roaming the site (‘community health team’) providing education, answering any client questions, and discussing how to motivate unvaccinated friends and family, 2 persons supporting check-out procedures including vaccine card preparations, and 2 site managers overseeing staff and logistics. The security and safety of clients and staff were extremely important considerations and were provided by Promotores and members of San Francisco’s Community Ambassadors program.

All site personnel were selected based on their desire to serve their community with respect and compassion; they received initial and ongoing training emphasizing the importance of kindness and helpfulness and that the “client” needs should be understood and respected. All site personnel were also provided basic education on key facts related to COVID-19 vaccinations, based on the principle that any community member might ask any staff member basic COVID-19 vaccine questions at any time point in the process, and such question provide important teaching opportunities. Daily morning staff meetings occurred throughout the implementation period, which provided opportunities to discuss ongoing successes and challenges as well as changing COVID-19 vaccine eligibility criteria and any associated necessary adaptations in strategies; they also served as an important opportunity to provide staff refresher trainings on client-centeredness and updated education as new knowledge related to COVID-19 vaccines became available.

##### Low-barrier registration and vaccination approach

COVID-19 vaccine registration was originally available on-site, but it was quickly moved to the nearby UeS testing site given high demand, long lines and resultant congestion. We initially only provided in-person registration to preferentially provide access to eligible community members for whom online registration, as required at the time for most other vaccination sites, presented a barrier to access. In-person appointment scheduling was available onsite outside of operational hours and served as an opportunity for community members to interact and converse with one another while awaiting scheduling. To remove barriers to registration and vaccine appointment check-in, especially for clients who may have had immigration fears or concerns, community members were able to self-attest to their eligibility. Clients were not required to show any form of identification, or to provide proof of residence, or healthcare insurance in order to be scheduled. Upon arriving at the vaccination site, clients were checked in and scheduled for their second vaccine dose. Following vaccination, clients waited for at least 15 minutes in the post-vaccination waiting area (**Figure 2d)**, where site staff provided them education on adverse side effects to monitor for and ensured that any questions were answered (**Table 1)**. Those clients who did not have insurance or a primary care provider were set up with a community health partner who offered low barrier care in the event that they had any health concerns beyond the standard expected side effects of vaccination. Upon request, clients were provided letters for employers to account for their time away from work. Clients were provided their proof of vaccination only at the end of this observation period.

#### Leveraging social networks to increase vaccine uptake (“Activate”)

In recognition of the important role that vaccinated individuals play in influencing COVID-19 vaccine knowledge, attitudes and beliefs (and ultimately vaccine uptake) among their friends, family members, and co-workers, we sought to empower clients to become “vaccine ambassadors (**Table 1)**.” During the post-vaccination waiting period, two dedicated staff members (who also provided post-vaccination education and answered any questions), shared their personal experiences, encouraged clients to reach out to members of their social network that had not yet been vaccinated and share their positive vaccination experiences and recommend that they too get vaccinated. This simple act serves multiple functions, including that the peer vaccine ambassador can provide COVID-19 vaccine education and debunk common myths and misconceptions, demonstrate good health-seeking behavior, serve as a credible source for vaccination experiences and also to provide social support to get vaccinated [21–23]. Site staff provided tips on how to handle difficult conversations and role-played different hypothetical scenarios with clients to bolster confidence. All vaccinated clients were provided fliers with UeS neighborhood vaccination site registration information that could be handed out to unvaccinated friends, family members and coworkers. The flyer included a phone number for peer referrals of the vaccinated clients to call with any questions related to COVID-19 vaccines or registration.

### Evaluation of the “Motivate, Vaccinate, and Activate” Strategy

#### RE-AIM evaluation measures

We evaluated the “Motivate, Vaccinate, and Activate” strategy using the Reach, Effectiveness, Adoption, Implementation and Maintenance (RE-AIM) framework. We chose the RE-AIM framework as it allowed us to evaluate both individual client-level outcomes (reach and effectiveness), as well as site and community-level outcomes (effectiveness, implementation, and maintenance) [24]. Furthermore, RE-AIM has recently been updated to include an explicit focus on equity and to address dynamic implementation contexts that may require adaptive strategies to maintain interventions over time [12]. This provided an enhanced framework to evaluate our implementation strategy, which sought to facilitate equitable vaccine access and uptake among Latinx community members and also have components that could be adapted to respond to rapidly changing community needs and public health guidance.

- **Reach:** We sought to reach and increase vaccine uptake among any Latinx adults and adolescents living in San Francisco, with an emphasis on the Mission District, as soon as they became eligible according SFDPH guidance. We conceptualized reach at two levels (proximal and distal). The proximal reach of our vaccination program included the number of individuals directly reached by each implementation strategy component. The distal reach included the number of individuals who were received a COVID-19 vaccination at the UeS neighborhood vaccination site. Because of the community-based design of our implementation strategy, it is difficult to measure proximal reach, e.g., the exact number of Latinx community members who were reached through the community mobilization and demand generation activities, as a result of simply passing by the community-located vaccination site, or through contact with peer vaccine ambassadors. However, because the UeS neighborhood vaccination site was outside of the formal healthcare system, these activities were necessary precursors for community members to become aware of, schedule, and receive a vaccination at the UeS neighborhood vaccination site. Therefore, our evaluation focused on distal reach, e.g., the number of individuals scheduled for vaccination and vaccinated at the UeS neighborhood site. In order to evaluate whether our strategy reached Latinx people (the priority population our strategy was tailored for and aimed to reach) we also evaluated the demographic and socioeconomic characteristics of individuals vaccinated at the neighborhood vaccination site as measures of representativeness. We also assessed measures of geographic coverage by estimating the proportion of all Mission Distract (zip code 94110) residents vaccinated overall and among Latinx individuals as well as the proportion of all vaccinated individuals in San Francisco reached by the UeS neighborhood site overall and among Latinx persons.
- **Effectiveness:** There is strong evidence, including robust population-level data, that demonstrates that the COVID-19 vaccine, once administered, is highly effective in reducing the risk of COVID-19 disease and transmission [25–28]. Therefore, measures of effectiveness associated with the multicomponent implementation strategy used indicators of behavior change, including the proportion of clients who said that they were able to get vaccinated more quickly had the neighborhood site not existed and the proportion of clients who stated that they were more likely to reach out to and recommend vaccination to their unvaccinated friends, family members and coworkers after their experiences at the UeS neighborhood vaccination site. We also evaluated the proportion of clients at the neighborhood vaccination site who completed their second vaccine dose [29], as this metric may reflect a number of aspects of fidelity to and acceptability of the “Motivate, Vaccinate, and Activate” strategy and is therefore a composite quality outcome measure.
- **Implementation:** Implementation outcomes assessed were fidelity to each of the implementation strategy components (Motivate, Vaccinate, and Activate) as designed and also the acceptability of the overall implementation strategy among community members vaccinated through the UeS Neighborhood site.
- **Maintenance:** We evaluated maintenance in two ways. We first assessed temporal trends in the number of individuals receiving their first COVID-19 vaccination – overall and according to both eligibility criteria and ethnicity. This provided insight into the extent to which the “Motivate, Vaccinate, and Activate” strategy was able to evolve over time and mobilize different types individuals as they became eligible, while also being able to consistently reach Latinx individuals throughout the implementation period. We also documented and characterized adaptations during the implementation period.

#### Data sources and Statistics

Several data sources informed the evaluation of the “Motivate, Vaccinate, and Activate” strategy. Programmatic UeS vaccination data informed reach (including basic demographic chacteristics), effectiveness and maintenance related-outcomes. SFDPH surveillance data was used to inform estimates of vaccination coverage [30]. Census data informed population estimates in the Mission District (zip code 94110) [14]. To better understand the characteristics of those being reached, the possible reach and effectiveness of the peer vaccine ambassador strategy component, and the acceptability of our strategy among clients served by the neighborhood vaccination site, we administered a structured survey between May 2, 2021 and May 19, 2021. The survey data also captured additional client information including household income, occupation, insurance status and primary care status. It was administered on-site among those in the waiting area following completion of their vaccination (either first or second dose). Fidelity to and adaptations made to the implementation strategy components were assessed, discussed and documented throughout the implementation period as part of daily meetings with UeS neighborhood vaccination site workers and weekly meetings with UeS leadership.

Data were administratively censored at May 19, 2021, corresponding to a 16-week evaluation period. Analyses were restricted to adolescents and adults 16 years of age and older. Simple descriptive statistics were used to characterize individuals - Fisher’s exact or chi-squared tests were applied, as appropriate. As we primarily aimed to reach Latinx community members through our “Motivate, Vaccinate, and Activate” Strategy, all outcomes were assessed overall and according to whether individuals identified as Latinx (e.g., Latinx versus not-Latinx).

## Results

### Reach

#### COVID-19 vaccine uptake

Overall, there were 12,103 unique individuals registered for a COVID-19 vaccine at the UeS neighborhood vaccination site between February 1 and May 19, 2021, of which 11,098 (91.7%) received at least one vaccine dose at the neighborhood site; the proportion of persons registered who received at least one vaccine dose at the neighborhood site did not differ according to age, sex or ethnicity **(Supplementary Table 1).** In total, 20,792 COVID-19 vaccine doses were administered to community members ≥16 years old during the evaluation period.

The characteristics of 11,098 individuals receiving at least one COVID-19 vaccination at the UeS neighborhood vaccination site are shown in **Table 2**. Vaccine recipients had a median age of 43 (IQR, 32-56) years, 53.9% were male, and 70.5% were Latinx, 7.7% were Asian, 2.4% were black and 14.1% were white; 50.7% and 14.3% were either a first- or second-generation immigrant, respectively. The majority of clients receiving a vaccine dose worked in front-facing retail jobs and 61.0% of individuals had an annual household income of less than $50,000 per year (**Table 2**). More than one-third (36.9%) of clients did not have health insurance and nearly half (46.3%) did not have an established primary care provider. Latinx clients were substantially more likely than non-Latinx clients to have an annual household income of less than $50,000 a year (76.1% vs. 33.5%), be a first-generation immigrant (60.2% vs. 30.1%), not have health insurance (47.3% vs. 16.0%), and not have access to primary healthcare services (62.4% vs. 36.2%) (**Table 2**).

**Table 2.** Baseline characteristics of clients vaccinated at the Unidos en Salud neighborhood vaccination site, overall and according to ethnicity.

|  | **Overall**<br>**(N=11,098)** | **Latinx**<br>**(n=7,809)** | **Not Latinx**<br>**(n=3,289)** |
| --- | --- | --- | --- |
| **Median age, IQR** | 43 (32-56) | 43 (32-55) | 43 (32-58) |
| **Age Category** |  |  |  |
| 16-30 | 2530 (22.8%) | 1814 (23.2%) | 716 (21.8%) |
| 31-50 | 4658 (42.0%) | 3353 (42.9%) | 1305 (39.7%) |
| 50-64 | 2617 (23.6%) | 1793 (23.0%) | 824 (25.1%) |
| 65 and older | 1293 (11.7%) | 849 (10.9%) | 444 (13.5%) |
| **Sex** |  |  |  |
| Female | 4926 (44.4%) | 3434 (44.0%) | 1492 (45.4%) |
| Male | 5978 (53.9%) | 4303 (55.1%) | 1675 (50.9%) |
| Non-binary/other | 194 (1.7%) | 72 (0.9%) | 122 (3.7%) |
| **Immigration classification<sup>^</sup>** |  |  |  |
| First-generation immigrant | 149 (50.7%) | 121 (60.2%) | 28 (30.1%) |
| Second-generation immigrant | 42 (14.3%) | 29 (14.4%) | 13 (14.0%) |
| Neither first- or second-generation immigrant | 103 (35.0%) | 51 (25.4%) | 52 (55.9%) |
| **Primary occupation<sup>^</sup>** |  |  |  |
| Food and beverage | 162 (16.6%) | 139 (21.3%) | 23 (7.10%) |
| Tradesperson, cleaning, personal services | 161 (16.5%) | 136 (20.8%) | 25 (7.7%) |
| Finance, sales and technology | 140 (14.3%) | 43 (6.6%) | 97 (29.9%) |
| Retired/homemaker | 41 (4.19%) | 33 (5.1%) | 8 (2.5%) |
| Unemployed | 101 (10.3%) | 68 (10.4%) | 33 (10.2%) |
| Education | 42 (4.3%) | 31 (4.7%) | 11 (3.4%) |
| Student | 109 (11.1%) | 84 (12.8%) | 25 (7.7%) |
| Healthcare | 22 (2.3%) | 11 (1.7%) | 11 (3.4%) |
| Other | 200 (20.4%) | 109 (16.7%) | 91 (28.1%) |
| **Annual household income<sup>^</sup>** |  |  |  |
| <\$50,000 per year | 536 (61.0%) | 431 (76.1%) | 105 (33.5%) |
| \$50,000-100,000 per year | 199 (22.6%) | 107 (18.9%) | 92 (29.4%) |
| >\$100,000 per year | 144 (16.4%) | 28 (5.0%) | 116 (37.1%) |
| **Health insurance<sup>^</sup>** |  |  |  |
| Yes | 618 (63.1%) | 344 (52.7%) | 274 (84.0%) |
| No | 361 (36.9%) | 309 (47.3%) | 52 (16.0%) |
| **Health insurance type (if insured)<sup>^</sup>** |  |  |  |
| Public | 181 (29.5%) | 145 (42.6%) | 36 (13.1%) |
| Private | 391 (63.7%) | 173 (50.9%) | 218 (79.6%) |
| Unsure | 42 (6.84%) | 22 (6.5%) | 20 (7.3%) |
| **Primary care provider<sup>^</sup>** |  |  |  |
| Yes | 451 (46.3%) | 245 (37.6%) | 206 (63.8%) |
| No | 523 (53.7%) | 406 (62.4%) | 117 (36.2%) |

**Note:** Age, sex, and ethnicity are drawn from programmatic data among all vaccinated clients between February 1 and May 19<sup>th</sup>, 2021 (n=11,098). <sup>^</sup>Represents the results of a survey among vaccinated clients aged ≥16 years old conducted after their first or second vaccine dose between May 2 and 19<sup>th</sup>, 2021 (n=997).

Next, we assessed COVID-19 vaccine coverage associated with the UeS Neighborhood vaccination site. Among all eligible individuals (≥16 years old) estimated to be living in the Mission District (zip code 94110), 5.7% (n=3,590/62,452) received at least one vaccine dose at the neighborhood site; this included 11.9% (n=2,484/20,859) of the estimated number of Latinx residents. Compared to the ethnic makeup of the Mission District, clients receiving at least one vaccine dose at the neighborhood vaccination site were far more likely to be Latinx (70.5% vs. 33.4%) and far less likely to be white (14.1% vs. 44.8%) (**Figure 3**). While the neighborhood site was based in the Mission District (zip code 94110), it had broad geographic reach, such that less than one-third (32.3%) of all vaccine recipients resided in the Mission District (**Figure 4)**; the vaccination site was accessed by a large number of predominantly Latinx individuals residing and working throughout San Francisco and the Bay Area (**Supplementary Table 2**). The neighborhood site also appeared to reach persons residing predominantly in the neighborhoods of San Francisco that have been the most impacted by COVID-19 (**Figure 4, Supplementary Figure 1, Supplementary Table 2).**

**Figure 3.**
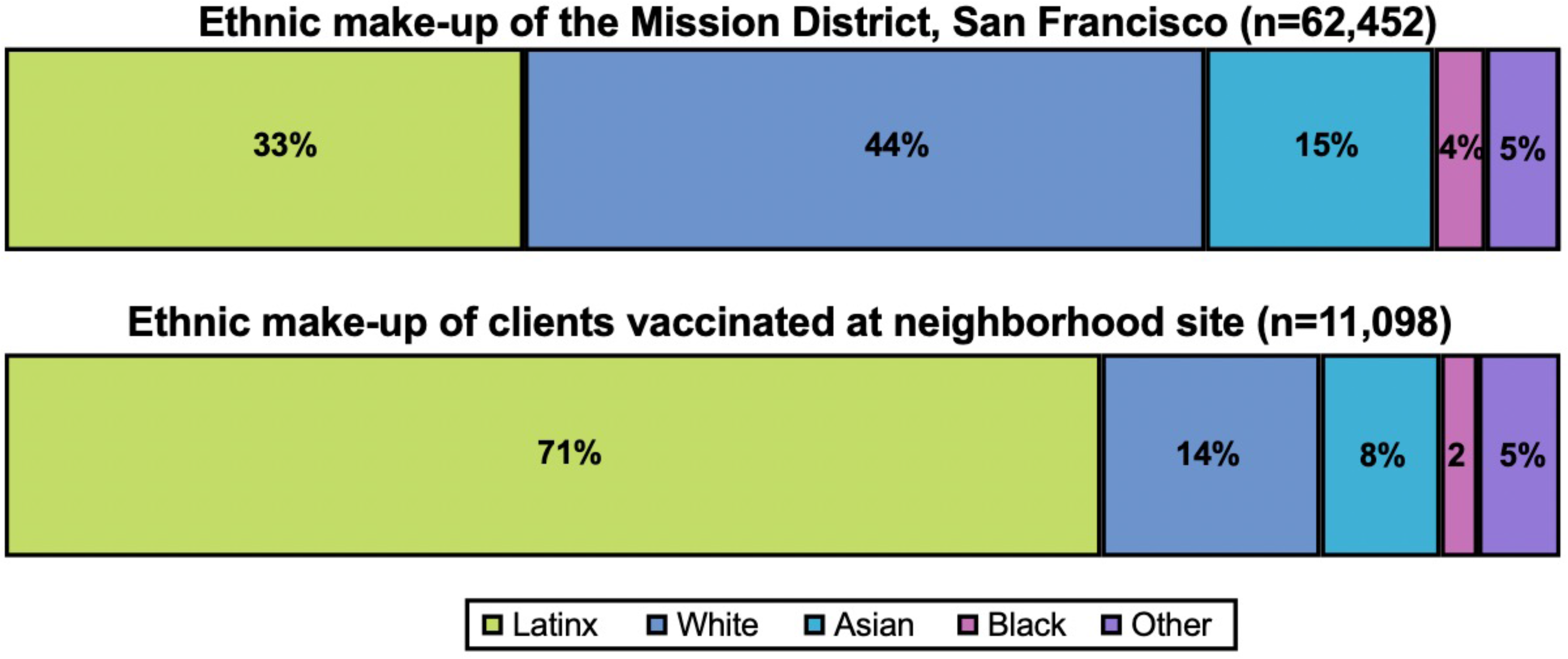
The ethnic composition of clients vaccinated at the Unidos en Salud neighborhood site (n=11,098) compared to the ethnic composition of the Mission District (n=62,452) among persons at least 16 years of age.

**Figure 4.**
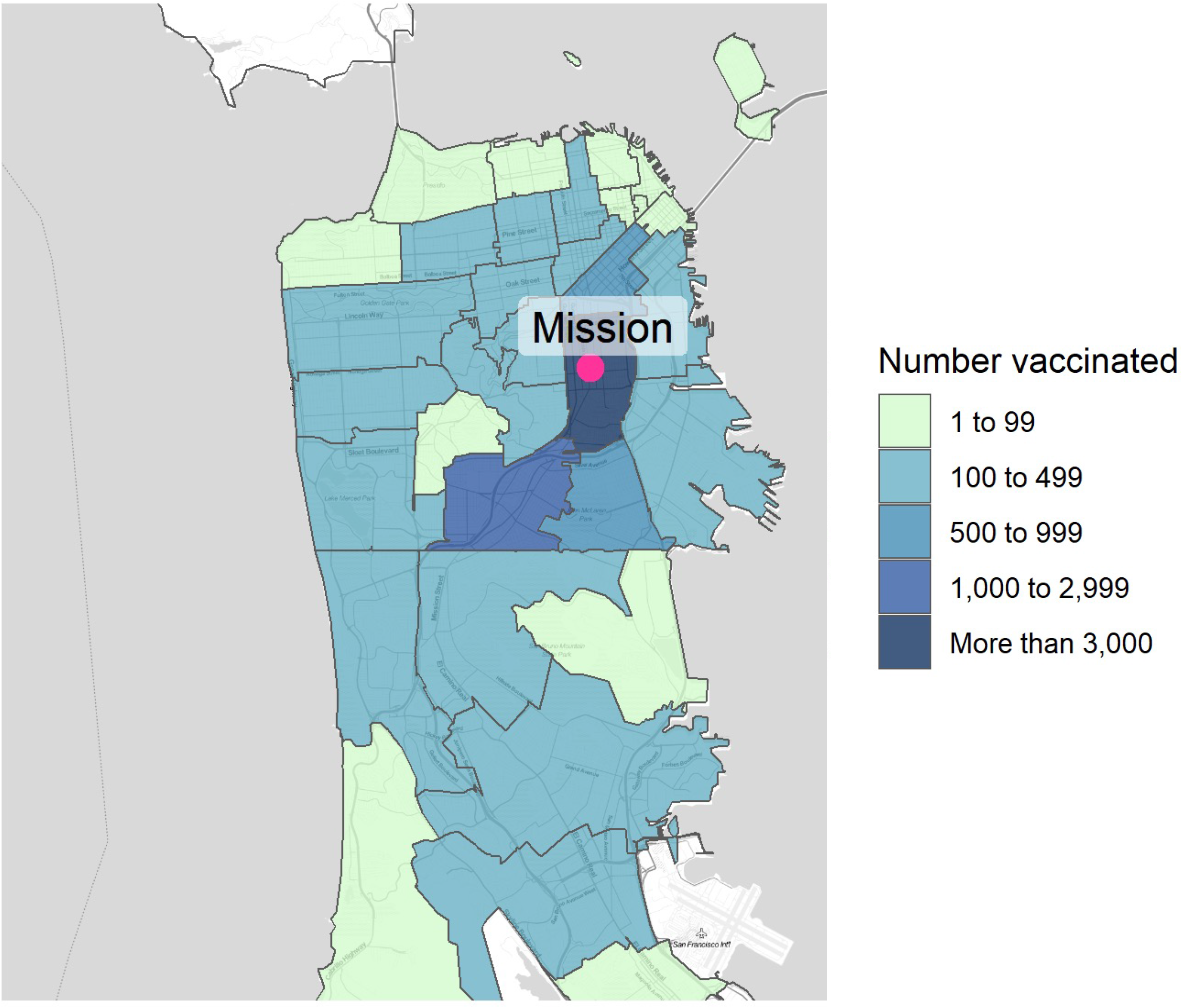
Map of San Francisco and the greater South Bay Area demonstrating the number of clients vaccinated at the Unidos en Salud neighborhood vaccination site according to their zip code of residence. The large majority of vaccinated clients living outside of San Francisco work in the Mission District.

#### Factors influencing clients to get vaccinated at neighborhood site

Of 3,597 clients offered to take a survey about their vaccination experiences, 997 (27.7%) completed the survey; compared to those who declined survey participation, survey respondents were slightly younger and less likely to be Latinx (**Supplementary Table 3).** Clients who were vaccinated at the UeS neighborhood vaccination site reported that they had heard about or became aware of the site in a number of different ways (**Table 3**). Clients most commonly found out about the site was from a friend, family member or co-worker (36.1%); a large number of clients also reported receiving a text invitation on their phone (21.0%), walking past the community-based site (17.8%) and receiving a direct referral from the nearby UeS COVID-19 testing site (11.4%). Clients less commonly cited having been made aware of the site through outreach from a community volunteer, or via a flyer, social media, or news sources (**Table 3**). Notably, the proportion of clients stating that they heard about the site through a friend or family member did not differ substantially by ethnicity (37.8% vs. 32.6%; **Table 3**). However, compared to non-Latinx clients, Latinx clients were more likely to report hearing about the site by directly passing by it in the neighborhood, and were less likely to have received a direct text invitation (**Table 3).**

**Table 3.** Factors influencing and motivating clients to get vaccinated at the at the Unidos en Salud neighborhood site.

|  | **Overall**<br>**(n=997)** | **Latinx**<br>**(n=669)** | **Not Latinx**<br>**(n=328)** | **P-value** |
| --- | --- | --- | --- | --- |
| **How clients heard about the site** |  |  |  |  |
| Told about it from a friend, family member, or co-worker | 352 (36.1%) | 248 (37.8%) | 104 (32.6%) | <0.001 |
| Received an invitation on their phone | 205 (21.0%) | 109 (16.6%) | 96 (30.1%) |  |
| Passed by the site | 174 (17.8%) | 144 (22.0%) | 30 (9.4%) |  |
| Referred from UeS testing site | 111 (11.4%) | 69 (10.5%) | 42 (13.2%) |  |
| Told about it by a community volunteer at their business or home | 57 (5.9%) | 35 (5.3%) | 22 (6.9%) |  |
| Saw a flyer for the site in the community | 23 (2.4%) | 13 (2.0%) | 10 (3.1%) |  |
| Saw information in the news | 9 (0.9%) | 7 (1.1%) | 2 (0.6%) |  |
| Saw information on social media | 15 (1.5%) | 8 (1.2%) | 7 (2.2%) |  |
| Other | 29 (3.0%) | 23 (3.5%) | 6 (1.9%) |  |
| **Reasons clients chose the site** |  |  |  |  |
| Located in their neighborhood | 251 (29.0%) | 170 (29.3%) | 81 (28.5%) | <0.001 |
| Scheduling was easy, efficient and convenient | 231 (26.7%) | 130 (22.4%) | 101 (35.6%) |  |
| Someone they trusted either invited them or suggested it to them | 156 (18.0%) | 111 (19.1%) | 45 (15.8%) |  |
| Had a positive interaction with the UeS of LTF staff/volunteers | 41 (4.7%) | 32 (5.5%) | 9 (3.2%) |  |
| Tried to get vaccinated somewhere else but was unsuccessful | 37 (4.3%) | 19 (3.3%) | 18 (6.3%) |  |
| They were not aware of other options | 46 (5.3%) | 37 (6.4%) | 9 (3.2%) |  |
| Staff was bilingual | 48 (5.6%) | 45 (7.8%) | 3 (1.1%) |  |
| Prior positive experience at UeS neighborhood testing site | 40 (4.6%) | 28 (4.8%) | 12 (4.2%) |  |
| Other | 15 (1.7%) | 9 (1.6%) | 6 (2.1%) |  |

**Note:** All data is drawn from a survey among vaccinated clients aged ≥16 years old conducted after their first or second vaccine dose between May 2 and 19<sup>th</sup>, 2021 (n=997).

Clients reported that their single most important reason for choosing to get vaccinated at the UeS neighborhood site was (1) because it was in their neighborhood (29.0%), (2) because scheduling was easy and convenient (26.7%) and (3) because someone they trusted had recommended it to them (18.0%) (**Table 3)**. Latinx clients were more likely to choose the site because of its bilingual staff compared non-Latinx clients but were less likely to cite the ease and efficiency of scheduling as an enabling factor (**Table 3)**.

### Implementation

#### Fidelity

Overall, we were able to deliver each of the components of the “Motivate, Vaccinate, and Activate” strategy as originally intended. As intended by design, the strategy was adapted in response to rapid evolving eligibility criteria and site capacity (**Table 4).** Most adaptations to the strategy occurred early on and were related to the “Vaccinate” component of the strategy. We aimed to provide timely vaccination to all community members who were eligible and wished to be vaccinated at the neighborhood site. While that was often possible, at times, peaks in demand exceeded our capacity to provide immediate vaccinations. In order to not delay vaccination among highly motivated community members, we partnered and worked closely with a local safety net hospital to extend the reach of our strategy by facilitating referrals for typically either same-day or next-day vaccination appointments. There were two key features of this adaption: (1) free transportation was provided to any referred client who needed it and (2) a UeS community team member went with referred clients or met them at the hospital to provide support and help navigate any additional barriers to getting vaccinated. While complete estimates are not available, more than 2,400 additional community members were directly referred and scheduled for vaccination through this strategy, including approximately 850 during the second week of March 2021. We believe that this was a key adaptation that was needed as to not undermine the overall effectiveness of our strategy, however, it was only required for less than four weeks.

**Table 4.** Adaptations to the “Motivate, Vaccinate and Activate” strategy components during the implementation period from February 1 through May 19, 2021.

| Strategy component | Original design / aim | Description of adaptation(s) made | Was adaptation planned or unplanned? | When was adaptation made? | Why was adaptation made? |
| --- | --- | --- | --- | --- | --- |
| **Motivate**<br>(Community mobilization and demand generation activities) | Multi-method outreach approach to mobilize community members and generate demand for COVID-19 vaccination. | • We initially focused on direct recruitment from CBO networks, those eligible on our testing lists, and in-person registration at the site (capitalizing on it being located at a busy transport hub).<br>• As eligibility expanded, we began undertaking direct sign ups at other sites (i.e., food hubs, grocery stores).<br>• We also began providing flyers/cards with QR codes to sign-up for vaccination at key locations (i.e., food hubs, school drop off/pick up)<br>• Later, we began promoting a direct sign-up link (e.g., via community signage, websites) for anyone to register. | Planned | Throughout the implementation period | • To address changing vaccine eligibility.<br>• To maintain demand for vaccination. |
| **Vaccinate**<br>(Community-based, low-barrier, client-centered, vaccination site) | Provide vaccinations 4 days a week without any unplanned closures in order to avoid the inconvenience of rescheduling among socioeconomically vulnerable individuals. | • Infrequently needed to close the site on a planned operating day | Unplanned | Throughout the implementation period | • To be responsive to events affecting local community (i.e., George Floyd verdict)<br>• No closures due to vaccine stock-out. |
|  | We aimed to provide evening hours to facilitate improved access to those who work. | • Unable to provide evening hours (site open 9am-4pm) | Unplanned | Early | • Given outdoor, community location, evening hours were not felt to be safe. |
|  | In-person scheduling co-located at the UeS neighborhood vaccination site in order to improve convenience. | • In-person scheduling moved to UeS neighborhood testing site. | Unplanned | Early | • To reduce congestion.<br>• To improve efficiency (many direct referrals from testing site). |
|  | Provide timely vaccination to all eligible community members who desired to be vaccinated at neighborhood site. | • Provided vaccination referrals to local safety net hospital.<br>• We worked closely with local hospital staff to utilize open same- and next-day appointments.<br>• Referred clients provided transport if needed<br>• Referred clients either escorted by or met at hospital by UeS site to provide support. | Unplanned | Early-middle | • At times demand outpaced timely appointments available.<br>• To get motivated community members vaccinated as soon as possible. |
|  | Only on-site, in-person registration offered in order to prioritize access for those without computer access/skills. | • On-line registration offered | Unplanned | Late | • To address changing vaccine eligibility.<br>• To maintain demand for vaccination. |

#### Acceptability

The UeS neighborhood site was highly acceptable to clients who were vaccinated there. Of 997 clients completing the survey, 98.6% stated that they would recommend the site to others; this did not differ between Latinx and non-Latinx clients **(Supplementary Table 4).** Clients were more likely to say that they would recommend the site to family members (82.1%) and friends (84.5%) than to co-workers (67.1%) (**Supplementary Table 4)**.

The features of the neighborhood vaccination site and their vaccination experience that clients stated that they liked the most were (1) the friendly and professional staff (40.3%), and (2) that the process was fast and efficient (32.8%) (**Supplementary Table 4**). Latinx clients were more likely to say that staff friendliness and professionalism was the site feature they liked the most about the neighborhood site, while non-Latinx clients were more likely to state they most liked the overall efficiency of the process (on average wait time was less than 5 minutes from check in) (**Figure 5a**). Clients stated that they liked and appreciated many additional features of the neighborhood vaccination site and preferences differed by ethnicity (**Figure 5b, Supplementary Table 4**). Latinx clients were more likely to state they liked that the staff was bilingual. Notably, more than a quarter of Latinx (26.3%) and non-Latinx clients (30.5%) reported that they liked that they did not need to show documentation of residency or proof of vaccine eligibility. Very few clients reported any aspects of their experience that they disliked. A few clients noted that they had experienced long wait times but were not bothered by it, while one client felt that the outdoor setting was not private enough. Several people responded that the site experience could be further improved by providing onsite toilets.

**Figure 5.**
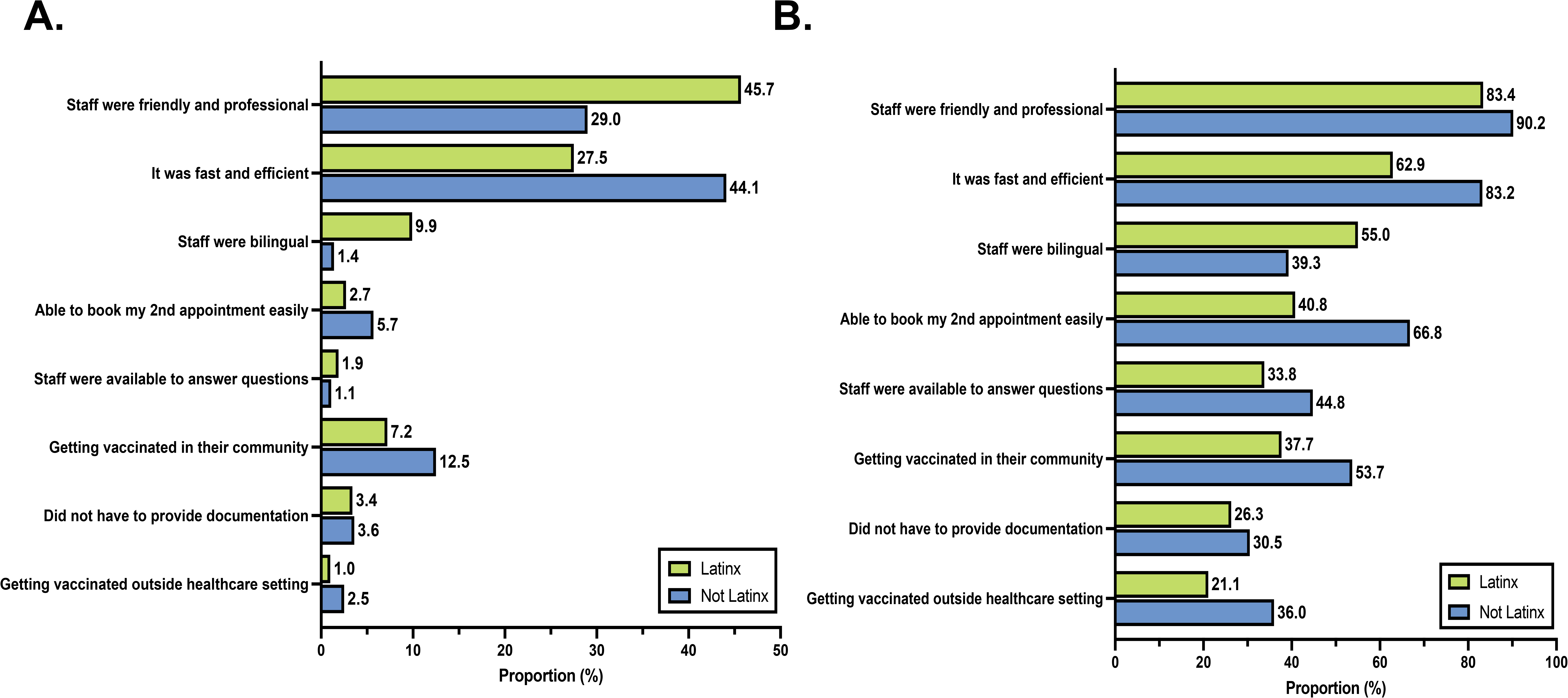
Features of the Unidos en Salud neighborhood vaccination site that clients said that they liked or appreciated stratified according to ethnicity. Panel A shows the features that clients liked the most (can choose only one) while Panel B shows all of the features that clients stated that they liked.

#### Maintenance

Several adaptations were made during the early implementation period, but there were very few subsequent adaptations and the “Motivate, Vaccinate, and Activate” strategy was delivered with high fidelity over time (**Table 4)**. An important feature that allowed us to continue to deliver our strategy with fidelity was that there was consistent staffing that helped facilitate group communication and cohesiveness.

As the eligibility for COVID-19 vaccination shifted over time in San Francisco, the “Motivate, Vaccinate, and Activate” strategy was able to continue to reach and facilitate vaccination of newly eligible community members (**Figure 6a**). Notably, despite evolution of the eligibility criteria over time, the large majority of clients reached throughout the entire implementation period were Latinx (**Figure 6b**). While the number of new individuals vaccinated in May 2021 declined (mirroring local and national trends) (**Figure 6a)**, the proportion of daily vaccinations that were among Latinx individuals increased (**Figure 6b**).

**Figure 6.**
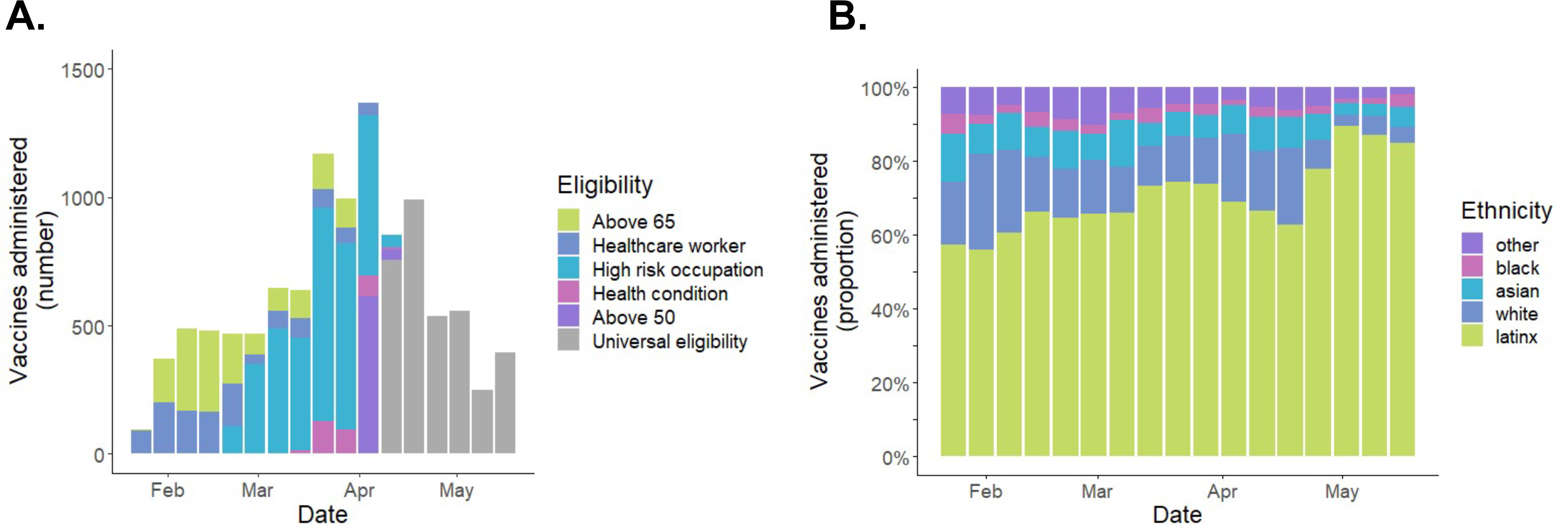
Temporal trends in vaccinations at the Unidos en Salud neighborhood vaccination site during the 16-week implementation period from February 1 to May 19, 2021. Panel A shows the number of vaccinations that were administered each week according to eligibility criteria indication. Panel B shows the proportion of all vaccinations each week that were administered to Latinx persons; this provides measure of how effectively Latinx individuals were reached throughout the entirety of the implementation period.

#### Effectiveness

##### Indicators of positive behavior change

There were 58.4% of clients that said they got vaccinated sooner than they otherwise would have had the neighborhood vaccination site not existed; this included 56.1% of Latinx clients and 63.2% of non-Latinx clients (**Table 5)**. After their experiences at the neighborhood vaccination site, 90.1% of clients said they were more likely to recommend getting vaccinated to family members, friends, and co-workers; this did not meaningfully differ by ethnicity (**Table 5).**

**Table 5.** Behavior change outcomes associated with the effectiveness of the “Motivate, Vaccinate and Activate” strategy.

|  | **Overall**<br>**(n=997)** | **Latinx**<br>**(n=669)** | **Not Latinx**<br>**(n=328)** | **P-value** |
| --- | --- | --- | --- | --- |
| **Had the neighborhood vaccination site not existed, when would you have been vaccinated?** |  |  |  |  |
| Later | 565 (58.4%) | 366 (56.1%) | 199 (63.2%) | 0.013 |
| About the same time | 335 (34.6%) | 231 (35.4%) | 104 (33.0%) |  |
| Earlier | 67 (6.9%) | 55 (8.4%) | 12 (3.8%) |  |
| **After your experience at the UeS site, were you more likely to recommend vaccination to family members, friends or co-workers?^** |  |  |  |  |
| Yes | 450 (89.1%) | 272 (90.1%) | 178 (87.7%) | 0.49 |
| No | 55 (10.9%) | 30 (9.9%) | 25 (12.3%) |  |
| **Since getting vaccinated at the UeS site, how many people have you reached out to get vaccinated?^** |  |  |  |  |
| 0 | 40 (9.0%) | 20 (7.4%) | 20 (11.4%) | 0.21 |
| 1-2 | 131 (29.4%) | 74 (27.3%) | 57 (32.6%) |  |
| 3-5 | 151 (33.9%) | 99 (36.5%) | 52 (29.7%) |  |
| 6-10 | 51 (11.4%) | 35 (12.9%) | 16 (9.1%) |  |
| >10 | 73 (16.4%) | 43 (15.9%) | 30 (17.1%) |  |
| **How many previously unvaccinated people are you aware of that got vaccinated after you recommended it to them?^** |  |  |  |  |
| 0 | 66 (17.0%) | 30 (12.5%) | 36 (24.3%) | 0.035 |
| 1-2 | 156 (40.2%) | 98 (40.8%) | 58 (39.2%) |  |
| 3-5 | 93 (24.0%) | 65 (27.1%) | 28 (18.9%) |  |
| 6-10 | 34 (8.8%) | 22 (9.2%) | 12 (8.1%) |  |
| >10 | 39 (10.1%) | 25 (10.4%) | 14 (9.5%) |  |

**Note:** All data is drawn from a survey among vaccinated clients aged ≥16 years old conducted after their first or second vaccine dose between May 2 and 19<sup>th</sup>, 2021 (n=997). ^ Responses limited to among participants completing the survey after their second vaccine dose (n=729).

Approximately 40% (40.3%) of clients said that they knew at least one person that had yet to be vaccinated; this was slightly higher among Latinx clients than non-Latinx clients (42.9% vs. 34.7%). Of clients who reported knowing unvaccinated individuals, 64.6% reported knowing 3 or more. Among clients who received both vaccine doses (n=729), 91.0% said that after their first vaccination experience, they personally reached out to at least one unvaccinated person they knew to recommend COVID-19 vaccination; Latinx clients were more likely than non-Latinx clients to reach out and recommend vaccination to 3 or more persons (65.3% vs. 55.9%; **Table 5**). Notably, 83.0% of clients stated that they were aware of at least 1 family member, friend or co-worker who got vaccinated as a result of their direct outreach; Latinx clients were more likely than non-Latinx persons to report that 3 or more persons got vaccinated a result of their influence (46.7% vs. 36.5%; **Table 5**).

Next, we evaluated the proportion of clients receiving their first COVID-19 vaccine dose at the UeS neighborhood site, who also completed their second vaccine dose. Among 9,305 clients with at least 4 weeks of follow-up time since their first vaccine dose, 9,152 (98.4%) completed their second dose; the proportion of clients completing both vaccine doses did not differ according to age, sex or ethnicity (**Supplementary Table 5**).

## Discussion

From the onset of the COVID-19 pandemic, there have been stark racial and ethnic disparities of populations infected and vaccinated, reflecting known health inequities in the United States [7, 8]. In the setting of a rapidly moving pandemic such as COVID-19, the challenge is how to overcome in a short time period the decades of disparate access to care and resulting mistrust among vulnerable populations in a way that improves health outcomes and can lead to sustained gains in health delivery. Via a community, academic and public health partnership (Unidos en Salud) we developed and evaluated a community-based, “Motivate, Vaccinate, and Activate” strategy. We sought to increase uptake of COVID-19 vaccination among Latinx persons and to activate clients to be community vaccine ambassadors within their social networks.

We effectively reached the target population—70% of more than 11,000 vaccine recipients during the evaluation period were Latinx, the majority of whom were first generation immigrants with a household income of less than $50,000 and without a primary care provider. The geographic reach extended to the Latinx community beyond the surrounding neighborhood to the Southeast sector of San Francisco, the area with the most COVID-19 cases in San Francisco [30]. The vaccination program was highly acceptable, with 99% of clients reporting they would recommend the site to others. The program was also highly effective, as 58% of people reported that they were vaccinated sooner because of the program, and among those who received both vaccine doses, over 90% of clients personally reached out to at least one person in their social network to recommend COVID-19 vaccination. Additionally, 98% of clients completed both vaccine doses, which is higher than early national estimates of 88% [29]. To our knowledge, this is the first evaluation of the implementation and effectiveness of a multicomponent community-based vaccination program designed to reach the Latinx community.

Our data suggest that efforts to address access and trust-related barriers underlie the effectiveness of the program. Access-related barriers drive a large portion of the COVID-19 vaccination disparities between Latinx and non-Latinx White people [10]. Our site was embedded in the San Francisco Mayor’s strategy to offer vaccination through multiple venues, including mass vaccination sites, health care and nursing home settings, pharmacies, and community sites. Local community sites can remove transportation barriers inherent to mass sites, mitigate issues of trust, and reach persons who are not actively connected with a formal health system. Despite all San Franciscan’s being eligible for health care, many vulnerable community members do not identify as having a primary physician or health insurance. Reasons may include the many administrative steps required for health care registration, language barriers, lack of outreach and navigation services on the part of health institutions and mistrust of government institutions based on past negative experiences. Clients at our vaccine site reported geographic convenience outside of a hospital setting and ease of registration (computer access not required) among their top reasons for choosing the neighborhood vaccine site. Time away from work for front-line workers is a major consideration for engagement in health care. The speed and efficiency of the experience was among the top three factors that people appreciated about the site.

Our survey results are consistent with recent national surveys that highlighted the importance of access-related barriers among Latinx persons who had not yet received the vaccine, with concerns about missing work, transport to the site, and information gaps about cost and impact on legal status [10]. Placing vaccine sites in central locations such as such as in Grand Central Station in New York, or trusted community sites such as churches has also yielded promising results [31–34]. While convenience was an important feature of the vaccine site, it alone is likely insufficient, and ensuring trust in the vaccine itself is the first step to getting people to come to the site.

In the “Motivate” component of our model, we used multiple approaches to address concerns voiced by Latinx persons, drawn both from prior work of Unidos en Salud in San Francisco and from national surveys, including safety, cost, eligibility, and effects on immigration status [9, 10]. Strategies to address these concerns included high-touch methods such as ‘door-to-door’ vaccine education and registration and mobilization by trusted community leaders via their social networks. We also employed less resource-intensive vaccine promotion strategies such as Spanish-language media, in which Unidos En Salud community leaders provided information and answered questions on COVID-19 vaccines. Our community team posted flyers in the neighborhood, including at local businesses, and handed out educational information about vaccines at our adjacent COVID-19 testing site (Table 1).

Our data highlight the importance of trusted messengers in the decision to come to the community vaccine site [23]. Nearly 20% of clients said that the most important factor in their decision to come to the vaccine site was because someone they trusted recommended it to them, as opposed to less than 5% who heard about the site through a flyer in the community or media campaigns. These data are consistent with prior data on the positive impact of door to- door outreach [35] and endorsements from trusted community members on increasing vaccination from influenza [36], childhood vaccines [37], and HPV vaccines [38]. Our findings are also consistent with a multicomponent intervention involving mobile clinics and religious leaders as vaccine ambassadors that led to high uptake among COVID-19 vaccines among Black people living in a community in Southern California [34]. Multi-pronged approaches to community-led education and outreach can increase trust in vaccine safety, effectiveness, and the healthcare system, and are fundamental to facilitating forward movement along the entire continuum of vaccine hesitancy [36–41].

Paramount to our strategy was to create a convenient, language-concordant, and welcoming vaccination site. The client experience at the site, including efficiency, and access to bilingual staff and health education in the post-vaccination area likely amplified trust in both the vaccination site and the vaccine itself. This was evidenced by the finding that 99% of vaccinated clients reported that they would recommend the site to their friends or family members, and that nearly two-thirds recommended COVID-19 vaccination to 3 or more people in their social network. Additionally, friendly and professional staff were the features Latinx clients liked most about the neighborhood vaccination site.

Peer-referrals and social network interventions can increase trust in marginalized communities and rapidly diffuse innovations. It was notable that even when San Francisco exceeded more than 75% coverage of COVID-19 vaccination among residents ≥16 years old, the social networks of people at our vaccine site still included large numbers of unvaccinated friends and family members. That our clients most frequently heard about the UeS neighborhood vaccination site from a friend or family member (36%) and more than 80% of vaccinated clients then positively influenced an unvaccinated person they knew to get vaccinated strongly supports the value of social network interventions in our setting and their potential to build trust and reach unvaccinated people. Our community health team, a bilingual bicultural team of community members, provided additional education about COVID-19 vaccines, testing, answered related questions and shared their own experiences about encouraging friends and loved ones to get vaccinated. We hypothesize that these positive interactions addressed information gaps and empowered people to become ‘vaccine ambassadors,’ and reach out to their unvaccinated family members, friends and co-workers. These findings build upon a growing body of literature demonstrating the effectiveness of social network interventions for positive health promotion, including prevention strategies for HIV (PrEP), STI and HIV testing, as well as other health behaviors and outcomes including smoking cessation, alcohol misuse, and improving hemoglobin A1c levels in persons with diabetes [42].

Although most factors influencing vaccination at our site were similar across race and ethnicity, Latinx compared to non-Latinx were more likely to report that bilingual staff were an important factor for choosing the site. This highlights the importance of language and cultural concordance throughout all stages from community outreach and mobilization, through vaccination to address structural barriers, information-gaps about vaccine eligibility, and perceptions that the COVID-19 vaccine costs money and can impact one’s immigration status. Additionally, requirements to provide documentation increases access-related barriers, especially for first generation immigrants and people who work informal jobs. Over 40% of Latinx persons in a national survey cited concern about having to provide documentation and 40% feared that the process would impact their legal status [10]. To address these concerns and to lower the barrier to vaccinations, we did not require identification or proof of vaccine eligibility at our vaccine site, and approximately one third of clients reported that they appreciated this feature. Removing the requirement for identification or proof of residence or employment should be considered in the design of low-barrier vaccine sites.

Our implementation strategy quickly adapted to changes in vaccine eligibility which created surges in demand over time. (Table 4). Initially vaccine demand exceeded supply. To address this need and not turn people away, we expanded our vaccine site to also include a vaccine navigation hub and become a gateway to a higher volume vaccine site at the safety-net hospital nearby. To facilitate access to larger vaccine sites, we arranged free transportation, helped schedule appointments on-site, and had our community workers accompany clients to the hospital site in order to overcome mistrust and fear of formal health care systems. Later, as supply exceeded demand, we shifted our mobilization strategy away from posting flyers in the community and harnessing Spanish language media, to more individualized ‘one-on-one’ discussions and also a focus on a social network-based approach as persons at our vaccine still had a large number of people in their network who remained unvaccinated. Financial and non-financial incentives can be effective in promoting vaccinations and adaptations that include incentives are worth further consideration and study [37,43,44].

The evaluation of our program has some limitations. Our methods underestimate the program’s reach, as we could not quantify the number of people who were influenced by our multifaceted, community-based demand generation activities, but who were vaccinated at a different site. Additionally, our reported reach does not include over 2,000 direct referrals to the vaccination site at the nearby county hospital. Secondly, the structured survey on clients’ experiences was only completed during the period of general eligibility, and experiences may have differed compared to the beginning of the program. This can also be seen as a strength, as the findings are more generalizable to the current vaccine landscape—where supply is greater than demand and all adults are eligible for the vaccine. As with most multi-component interventions, we are unable to fully disentangle the relative effects of the different components and subcomponents of the overall strategy. There are also some limitations to our measurements of effectiveness; though a high proportion of people reported that someone they referred received a vaccine, we could not measure this directly. However, even if the peer referral did not result in a vaccination, it is likely that the referral served as a nudge further down the continuum towards vaccine confidence.

In conclusion, our “Motivate, Vaccinate, and Activate” vaccine promotion strategy reached a high proportion of Latinx residents in San Francisco. We attribute the success of the program to demand generation through trusted messengers and social networks, multi-faceted and adaptable mobilization strategies, and a convenient and welcoming neighborhood vaccine site. Our Unidos en Salud community, academic, and public health partnership and co-design was fundamental to the program and cannot be underestimated. Though this program was geared towards addressing the specific barriers and needs of the Latinx community in San Francisco, the fundamental pillars of this program can be adapted to other local contexts.

## Data Availability

All relevant data are within the manuscript and its Supporting Information files.

## Acknowledgements

We would like to thank the community members who participated in this initiative and the many vaccination site staff, community ambassadors, and volunteers. We thank the Chan Zuckerberg Initiative, Supervisor Hillary Ronen, Mayor London Breed, The San Francisco Department of Public Health and Dr. Grant Colfax, Dr. Naveena Bobba, Dr. Susan Phillips, Dr. Mary Mercer, Emily Reingold, and Dr. Ellen Chen. We gratefully acknowledge the San Francisco Latino Task Force, Bay Area Phlebotomy and Laboratory Services (BayPLS), Stacy Powers and BRAVA for Women in the Arts, Bevan Dufty and the BART team, PrimaryBio COVID-19 Testing Platform, Dr. Rachel Stern, Phillipa Doyle, and Ricardo Duarte from Zuckerberg San Francisco General Hospital, Dr. Jessica Briggs and Dr. Susa Coffey from UCSF, and Dara Fonseca, Jocelin Payan, and Jack Fukushima from Unidos en Salud. We also would like to thank Mike Kai Chen for the use of his photographs in this manuscript.

---

**Supplementary Table 1.** Characteristics of individuals who were registered for vaccination at the Unidos en Salud vaccination site between February 1 and May 19, 2021, according to whether they did or did not receive at least one vaccine dose at the Unidos en Salud vaccination site.

|  | **Registered for vaccination at UeS site (n=12,103)** | **Received at least one vaccine dose at UeS site (n=11,098)** | **Did not receive at least one vaccine dose at UeS site (n=1,005)** |
| --- | --- | --- | --- |
| **Median age, IQR** | 43 (32-56) | 43 (32-56) | 42 (30-54) |
| **Age Category** |  |  |  |
| 16-30 | 2796 (100%) | 2530 (90.5%) | 266 (9.5%) |
| 31-50 | 5064 (100%) | 4658 (92.0%) | 406 (8.0%) |
| 50-64 | 2850 (100%) | 2617 (91.8%) | 233 (8.2%) |
| 65 and older | 1393 (100%) | 1293 (92.8%) | 100 (7.2%) |
| **Sex** |  |  |  |
| Male | 5423 (100%) | 4926 (90.8%) | 497 (9.2%) |
| Female | 6469 (100%) | 5978 (92.4%) | 491 (7.6%) |
| Non-binary/other | 211 (100%) | 194 (91.9%) | 17 (8.1%) |
| **Ethnicity** |  |  |  |
| Latinx | 8458 (100%) | 7809 (92.3%) | 649 (7.7%) |
| White | 1716 (100%) | 1581 (92.1%) | 135 (7.9%) |
| Asian | 959 (100%) | 852 (88.8%) | 107 (11.3%) |
| Black | 313 (100%) | 269 (85.9%) | 44 (14.1%) |
| Other | 657 (100%) | 587 (89.3%) | 70 (10.7%) |

Note: proportions represent row percentages.

**Supplementary Table 2.**
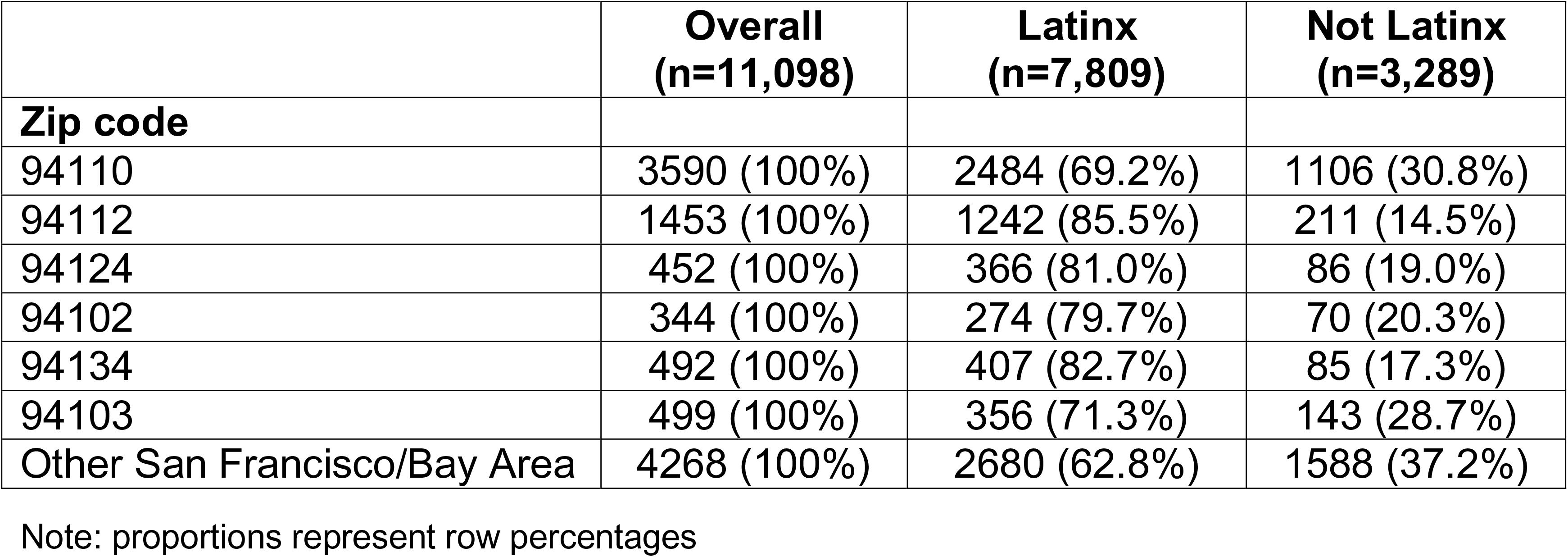
Geographic residence of clients receiving at least one vaccine dose at the Unidos en Salud neighborhood vaccination site between February 1 and May 19, 2021 according to zip code.

**Supplementary Table 3.** Characteristics of individuals who were offered participation in the on-site post-vaccination survey, according to whether they did or did not agree to complete the survey. All surveys were completed between May 2 and May 19^th^, 2021.

|  | **Offered**<br>**survey**<br>**(n=3,597)** | **Completed**<br>**survey**<br>**(n=997)** | **Declined**<br>**survey**<br>**(n=2,540)** |
| --- | --- | --- | --- |
| **Median age, IQR** | 37 (27-48) | 35 (26-46) | 38 (28-49) |
| **Age Category** |  |  |  |
| 16-30 | 1154 (32.6%) | 384 (38.5%) | 770 (30.3%) |
| 31-50 | 1676 (47.4%) | 430 (43.1%) | 1246 (49.1%) |
| 50-64 | 580 (16.4%) | 163 (16.3%) | 417 (16.4%) |
| 65 and older | 127 (3.6%) | 20 (2.0%) | 107 (4.2%) |
| **Sex** |  |  |  |
| Male | 2130 (60.2%) | 575 (57.7%) | 1555 (61.2%) |
| Female | 1359 (38.4%) | 409 (41.0%) | 950 (37.4%) |
| Non-binary/other | 48 (1.36%) | 13 (1.3%) | 35 (1.38%) |
| **Ethnicity** |  |  |  |
| Latinx | 2632 (74.4%) | 669 (67.1%) | 1963 (77.3%) |
| White | 427 (12.1%) | 163 (16.3%) | 264 (10.4%) |
| Asian | 242 (6.8%) | 83 (8.3%) | 159 (6.3%) |
| Black | 80 (2.3%) | 25 (2.5%) | 55 (2.2%) |
| Other | 156 (4.4%) | 57 (5.7%) | 99 (3.9%) |

**Supplementary Table 4.**
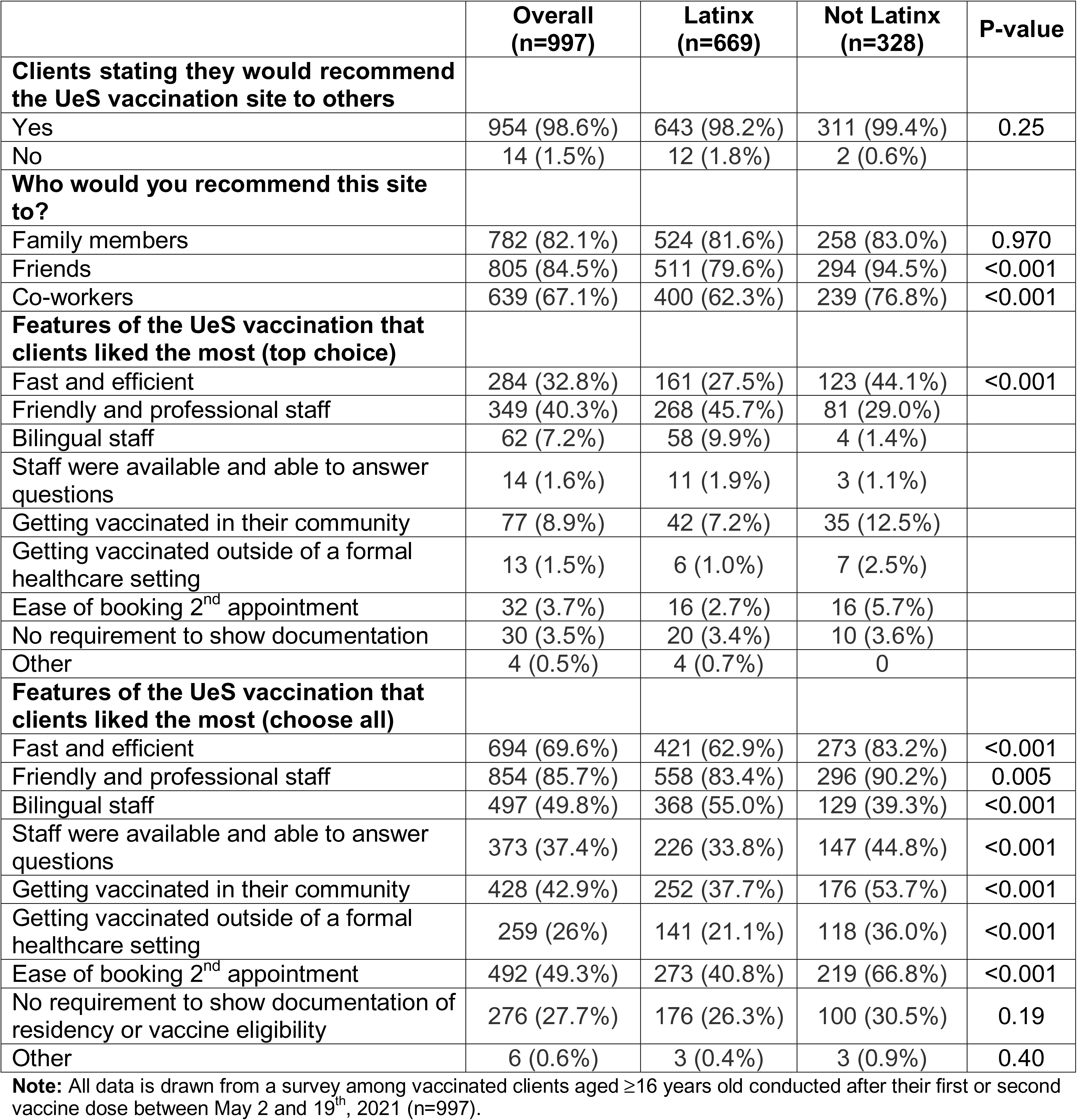
Acceptability measures associated with the Unidos en Salud vaccination site.

|  | **Overall**<br>**(n=997)** | **Latinx**<br>**(n=669)** | **Not Latinx**<br>**(n=328)** | **P-value** |
| --- | --- | --- | --- | --- |
| **Clients stating they would recommend the UeS vaccination site to others** |  |  |  |  |
| Yes | 954 (98.6%) | 643 (98.2%) | 311 (99.4%) | 0.25 |
| No | 14 (1.5%) | 12 (1.8%) | 2 (0.6%) |  |
| **Who would you recommend this site to?** |  |  |  |  |
| Family members | 782 (82.1%) | 524 (81.6%) | 258 (83.0%) | 0.970 |
| Friends | 805 (84.5%) | 511 (79.6%) | 294 (94.5%) | <0.001 |
| Co-workers | 639 (67.1%) | 400 (62.3%) | 239 (76.8%) | <0.001 |
| **Features of the UeS vaccination that clients liked the most (top choice)** |  |  |  |  |
| Fast and efficient | 284 (32.8%) | 161 (27.5%) | 123 (44.1%) | <0.001 |
| Friendly and professional staff | 349 (40.3%) | 268 (45.7%) | 81 (29.0%) |  |
| Bilingual staff | 62 (7.2%) | 58 (9.9%) | 4 (1.4%) |  |
| Staff were available and able to answer questions | 14 (1.6%) | 11 (1.9%) | 3 (1.1%) |  |
| Getting vaccinated in their community | 77 (8.9%) | 42 (7.2%) | 35 (12.5%) |  |
| Getting vaccinated outside of a formal healthcare setting | 13 (1.5%) | 6 (1.0%) | 7 (2.5%) |  |
| Ease of booking 2<sup>nd</sup> appointment | 32 (3.7%) | 16 (2.7%) | 16 (5.7%) |  |
| No requirement to show documentation | 30 (3.5%) | 20 (3.4%) | 10 (3.6%) |  |
| Other | 4 (0.5%) | 4 (0.7%) | 0 |  |
| **Features of the UeS vaccination that clients liked the most (choose all)** |  |  |  |  |
| Fast and efficient | 694 (69.6%) | 421 (62.9%) | 273 (83.2%) | <0.001 |
| Friendly and professional staff | 854 (85.7%) | 558 (83.4%) | 296 (90.2%) | 0.005 |
| Bilingual staff | 497 (49.8%) | 368 (55.0%) | 129 (39.3%) | <0.001 |
| Staff were available and able to answer questions | 373 (37.4%) | 226 (33.8%) | 147 (44.8%) | <0.001 |
| Getting vaccinated in their community | 428 (42.9%) | 252 (37.7%) | 176 (53.7%) | <0.001 |
| Getting vaccinated outside of a formal healthcare setting | 259 (26%) | 141 (21.1%) | 118 (36.0%) | <0.001 |
| Ease of booking 2<sup>nd</sup> appointment | 492 (49.3%) | 273 (40.8%) | 219 (66.8%) | <0.001 |
| No requirement to show documentation of residency or vaccine eligibility | 276 (27.7%) | 176 (26.3%) | 100 (30.5%) | 0.19 |
| Other | 6 (0.6%) | 3 (0.4%) | 3 (0.9%) | 0.40 |

**Note:** All data is drawn from a survey among vaccinated clients aged ≥16 years old conducted after their first or second vaccine dose between May 2 and 19<sup>th</sup>, 2021 (n=997).

**Supplementary Table 5.** Characteristics of clients receiving at least one vaccine dose at the Unidos en Salud vaccination site between February 1 and April 21, 2021 according to whether they completed both vaccine doses.

|  | **Total clients**<br>**receiving at**<br>**least one**<br>**dose**<br>**(n=9,305)** | **Completed**<br>**both doses**<br>**(n=9,152)** | **Did not**<br>**complete**<br>**both doses**<br>**(n=153)** |
| --- | --- | --- | --- |
| **Age Category** |  |  |  |
| 16-30 | 1900 (100%) | 1865 (98.2%) | 35 (1.8%) |
| 31-50 | 3822 (100%) | 3752 (98.2%) | 70 (1.8%) |
| 50-64 | 2353 (100%) | 2321 (98.6%) | 32 (1.4%) |
| 65 and older | 1230 (100%) | 1214 (98.8%) | 16 (1.2%) |
| **Sex** |  |  |  |
| Male | 4272 (100%) | 4221 (98.8%) | 51 (1.2%) |
| Female | 4851 (100%) | 4755 (97.8%) | 96 (2.2%) |
| Non-binary/other | 182 (100%) | 176 (96.7%) | 6 (3.3%) |
| **Ethnicity** |  |  |  |
| Latinx | 6313 (100%) | 6207 (98.3%) | 106 (1.7%) |
| White | 1476 (100%) | 1453 (98.4%) | 23 (1.6%) |
| Asian | 760 (100%) | 751 (98.8%) | 9 (1.2%) |
| Black | 230 (100%) | 227 (98.7%) | 3 (1.3%) |
| Other | 526 (100%) | 514 (97.7%) | 12 (2.3%) |

Note: Analysis is censored after April 21, 2021 in order to allow all clients equal opportunity to accrue 4 weeks of follow-up time after their first vaccine dose at the neighborhood vaccination site. There were 1,780 people excluded who received their 1<sup>st</sup> dose after April 21, 2021 and 13 excluded who received only their second dose at the neighborhood site.

**Supplementary Figure 1.**
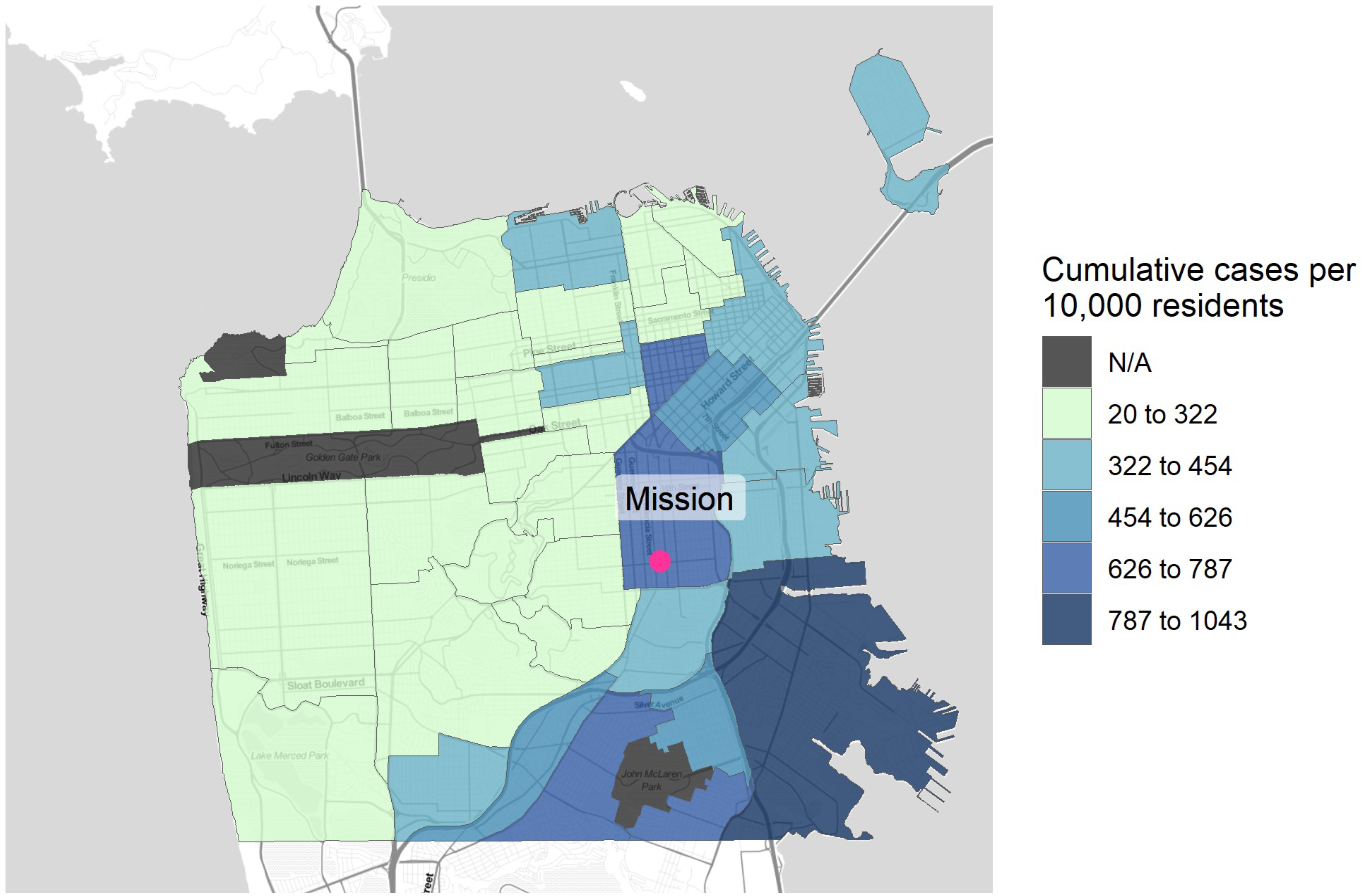
Map of San Francisco according to cumulative COVID-19 cases (rate per 10,000 residents) according to neighborhood. Darker blue shading indicates a higher cumulative prevalence of COVID-19 in a given neighborhood. The pink dot indicates the location of the Unidos en Salud neighborhood vaccination site.

## References

1. Chamie G, Marquez C, Crawford E, Peng J, Petersen M, Schwab D, et al. SARS-CoV-2 Community Transmission disproportionately affects Latinx population during Shelter-in-Place in San Francisco. Clin Infect Dis. 2020; ciaa1234-. doi:10.1093/cid/ciaa1234

2. Gil RM, Marcelin JR, Zuniga-Blanco B, Marquez C, Mathew T, Piggott DA. COVID-19 Pandemic: Disparate Health Impact on the Hispanic/Latinx Population in the United States. J Infect Dis. 2020; jiaa474-. doi:10.1093/infdis/jiaa474

3. Kullar R, Marcelin JR, Swartz TH, Piggott DA, Gil RM, Mathew TA, et al. Racial Disparity of Coronavirus Disease 2019 (COVID-19) in African American Communities. J Infect Dis. 2020;222: jiaa372-. doi:10.1093/infdis/jiaa372

4. Rodriguez-Diaz CE, Guilamo-Ramos V, Mena L, Hall E, Honermann B, Crowley JS, et al. Risk for COVID-19 infection and death among Latinos in the United States: examining heterogeneity in transmission dynamics. Ann Epidemiol. 2020;52: 46–53.e2. doi:10.1016/j.annepidem.2020.07.007

5. Bibbins-Domingo K, Petersen M, Havlir D. Taking Vaccine to Where the Virus Is—Equity and Effectiveness in Coronavirus Vaccinations. Jama Heal Forum. 2021;2: e210213. doi:10.1001/jamahealthforum.2021.0213

6. Hooper MW, Nápoles AM, Pérez-Stable EJ. No Populations Left Behind: Vaccine Hesitancy and Equitable Diffusion of Effective COVID-19 Vaccines. J Gen Intern Med. 2021; 1–4. doi:10.1007/s11606-021-06698-5

7. Ndugga N, Pham O, Hill L, Artiga S, Alam R, Parker N. Latest Data on COVID-19 Vaccinations Race/Ethnicity. Available at: https://www.kff.org/coronavirus-covid-19/issue-brief/latest-data-on-covid-19-vaccinations-race-ethnicity/. Accessed May 20, 2021.

8. California Department of Public Health. Vaccination progress data - Coronavirus COVID-19 Response. Available at: https://covid19.ca.gov/vaccination-progress-data/. Accessed May 20, 2021.

9. Peng J, Marquez C, Rubio L, Chamie G, Jones D, Jacobo J, et al. High likelihood of accepting COVID-19 vaccine in a Latinx community at high SARS-CoV2 risk in San Francisco. Open Forum Infect Dis. 2021; ofab202-. doi:10.1093/ofid/ofab202

10. Kaiser Family Foundation. KFF Covid-19 vaccine monitor: April 2021. Available at: https://www.kff.org/coronavirus-covid-19/poll-finding/kff-covid-19-vaccine-monitor-april-2021/. Accessed May 21, 2021.

11. Williams DR, Cooper LA. Reducing Racial Inequities in Health: Using What We Already Know to Take Action. Int J Environ Res Pu. 2019;16: 606. doi:10.3390/ijerph16040606

12. Shelton RC, Chambers DA, Glasgow RE. An Extension of RE-AIM to Enhance Sustainability: Addressing Dynamic Context and Promoting Health Equity Over Time. Frontiers Public Heal. 2020;8: 134. doi:10.3389/fpubh.2020.00134

13. Urban Displacement Project. Rising Housing Costs and Re-Segregation in San Francisco. 2018 Sep. Available: https://www.urbandisplacement.org/sites/default/files/images/sf_final.pdf. Accessed May 21, 2021.

14. Census Reporter. ZIP Code Tabulation Area 94110, San Francisco, CA. Available at: https://censusreporter.org/profiles/86000US94110-94110/. Accessed May 21, 2021.

15. Pilarowski G, Lebel P, Sunshine S, Liu J, Crawford E, Marquez C, et al. Performance characteristics of a rapid SARS-CoV-2 antigen detection assay at a public plaza testing site in San Francisco. J Infect Dis. 2021;223: jiaa802-. doi:10.1093/infdis/jiaa802

16. Kerkhoff AD, Sachdev D, Mizany S, Rojas S, Gandhi M, Peng J, et al. Evaluation of a novel community-based COVID-19 ‘Test-to-Care’ model for low-income populations. Plos One. 2020;15: e0239400. doi:10.1371/journal.pone.0239400

17. Rubio LA, Peng J, Rojas S, Rojas S, Crawford E, Black D, et al. The COVID-19 Symptom to Isolation Cascade in a Latinx Community: A Call to Action. Open Forum Infect Dis. 2021;8: ofab023. doi:10.1093/ofid/ofab023

18. Fields J, Gutierrez RJ, Marquez C, Rhoads K, Kushel M, Fernandez A, et al. Community-Academic Partnerships to Address COVID-19 Inequities: Lessons from the San Francisco Bay Area. NEJM Catal. 2021. In press.

19. The San Francisco Latino Task Force on COVID-19. Available at: https://www.ltfrespuestalatina.com/. Accessed May 21, 2021.

20. Porter CM. Revisiting Precede–Proceed: A leading model for ecological and ethical health promotion. Health Educ J. 2016;75: 753–764. doi:10.1177/0017896915619645

21. Funnell MM. Peer-based behavioural strategies to improve chronic disease self-management and clinical outcomes: evidence, logistics, evaluation considerations and needs for future research. Fam Pract. 2010;27: i17–i22. doi:10.1093/fampra/cmp027

22. Fisher EB, Ballesteros J, Bhushan N, Coufal MM, Kowitt SD, McDonough AM, et al. Key Features of Peer Support In Chronic Disease Prevention And Management. Health Affair. 2017;34: 1523–1530. doi:10.1377/hlthaff.2015.0365

23. National Academies of Sciences and Engineering Medicine. Strategies for Building Confidence in the COVID-19 Vaccines. Available at: https://www.nap.edu/catalog/26068/strategies-for-building-confidence-in-the-covid-19-vaccines. Accessed May 21, 2021.

24. Glasgow RE, Harden SM, Gaglio B, Rabin B, Smith ML, Porter GC, et al. RE-AIM Planning and Evaluation Framework: Adapting to New Science and Practice With a 20-Year Review. Frontiers Public Heal. 2019;7: 64. doi:10.3389/fpubh.2019.00064

25. Polack FP, Thomas SJ, Kitchin N, Absalon J, Gurtman A, Lockhart S, et al. Safety and Efficacy of the BNT162b2 mRNA Covid-19 Vaccine. New Engl J Med. 2020;383: 2603–2615. doi:10.1056/nejmoa2034577

26. Baden LR, Sahly HME, Essink B, Kotloff K, Frey S, Novak R, et al. Efficacy and Safety of the mRNA-1273 SARS-CoV-2 Vaccine. New Engl J Med. 2020;384: 403–416. doi:10.1056/nejmoa2035389

27. Haas EJ, Angulo FJ, McLaughlin JM, Anis E, Singer SR, Khan F, et al. Impact and effectiveness of mRNA BNT162b2 vaccine against SARS-CoV-2 infections and COVID-19 cases, hospitalisations, and deaths following a nationwide vaccination campaign in Israel: an observational study using national surveillance data. Lancet. 2021;397: 1819–1829. doi:10.1016/s0140-6736(21)00947-8

28. Benenson S, Oster Y, Cohen MJ, Nir-Paz R. BNT162b2 mRNA Covid-19 Vaccine Effectiveness among Health Care Workers. New Engl J Med. 2021;384: 1775–1777. doi:10.1056/nejmc2101951

29. Kriss JL, Reynolds LE, Wang A, Stokley S, Cole MM, Harris LQ, et al. COVID-19 Vaccine Second-Dose Completion and Interval Between First and Second Doses Among Vaccinated Persons — United States, December 14, 2020−February 14, 2021. Morbidity Mortal Wkly Rep. 2021;70: 389–395. doi:10.15585/mmwr.mm7011e2

30. San Francisco Department of Public Health: COVID-19 Data and Reports. Available at: https://data.sfgov.org/stories/s/fjki-2fab. Accessed May 23, 2021.

31. Cabanatuan M. Line around the block for COVID vaccinations in East Oakland. San Francisco Chronicle. 4 Apr 2021. Available: https://www.sfchronicle.com/local/article/Line-around-the-block-for-COVID-shots-in-East-16075137.php

32. Zaveri M. New York will offer vaccines at some subway stops through a pilot program. New York TImes. 10 May 2021. Available: https://www.nytimes.com/2021/05/10/nyregion/nyc-vaccine-subway.html

33. Weiland N. In New Vaccination Push, Biden Leans on His ‘Community Corps.’ New York Times. 16 May 2021. Available: https://www.nytimes.com/2021/05/16/us/politics/coronavirus-vaccination.html

34. Abdul-Mutakabbir JC, Casey S, Jews V, King A, Simmons K, Hogue MD, et al. A three-tiered approach to address barriers to COVID-19 vaccine delivery in the Black community. Lancet Global Heal. 2021;9: e749–e750. doi:10.1016/s2214-109x(21)00099-1

35. Centers for Disease Control and Prevention. Vaccine-preventable diseases: improving vaccination coverage in children, adolescents, and adults. A report on recommendations from the Task Force on Community Preventive Services. Morb Mortal Wkly Rep.1999;48:1–15.

36. Peterson P, McNabb P, Maddali SR, Heath J, Santibañez S. Engaging Communities to Reach Immigrant and Minority Populations: The Minnesota Immunization Networking Initiative (MINI), 2006-2017. Public Health Rep. 2019;134: 241–248. doi:10.1177/0033354919834579

37. Banerjee AV, Duflo E, Glennerster R, Kothari D. Improving immunisation coverage in rural India: clustered randomised controlled evaluation of immunisation campaigns with and without incentives. Bmj. 2010;340: c2220. doi:10.1136/bmj.c2220

38. Gottvall M, Tydén T, Höglund AT, Larsson M. Knowledge of human papillomavirus among high school students can be increased by an educational intervention. Int J Std Aids. 2010;21: 558–562. doi:10.1258/ijsa.2010.010063

39. Jarrett C, Wilson R, O’Leary M, Eckersberger E, Larson HJ, Hesitancy the SWG on V. Strategies for addressing vaccine hesitancy – A systematic review. Vaccine. 2015;33: 4180–4190. doi:10.1016/j.vaccine.2015.04.040

40. MacDonald NE, Hesitancy the SWG on V. Vaccine hesitancy: Definition, scope and determinants. Vaccine. 2015;33: 4161–4164. doi:10.1016/j.vaccine.2015.04.036

41. Force TAHCT, Spleen AM, Kluhsman BC, Clark AD, Dignan MB, Lengerich EJ. An Increase in HPV-Related Knowledge and Vaccination Intent Among Parental and Non-parental Caregivers of Adolescent Girls, Age 9–17 Years, in Appalachian Pennsylvania. J Cancer Educ. 2012;27: 312–319. doi:10.1007/s13187-011-0294-z

42. Hunter RF, Haye K de la, Murray JM, Badham J, Valente TW, Clarke M, et al. Social network interventions for health behaviours and outcomes: A systematic review and meta-analysis. PLOS Med. 2019;16: e1002890. doi:10.1371/journal.pmed.1002890

43. Vavreck L. $100 as Incentive to Get a Shot? Experiment Suggests It Can Pay Off. New York Times. 4 Mar 2021. Available: https://www.nytimes.com/2021/05/04/upshot/vaccine-incentive-experiment.html

44. McNamara D. Winning Idea: Ohio Vaccine Lottery Shows Some Incentives May Work. Medscape Medical News. 18 May 2021.

